# Spatial distribution of spinal cord fMRI activity with electrocutaneous stimulation

**DOI:** 10.64898/2026.02.26.26347215

**Authors:** Sandrine Bédard, Merve Kaptan, Teresa Indriolo, Christine SW Law, Dario Pfyffer, Lindsay Lee, John Ratliff, Serena Hu, Suzanne Tharin, Zachary A. Smith, Gary H. Glover, Sean Mackey, Julien Cohen-Adad, Kenneth A Weber

## Abstract

Sensory organization at the spinal segment level is commonly inferred from dermatomal maps that assume a fixed correspondence between cutaneous regions and spinal segments. However, based on the complexities of spinal neuroanatomy and neurophysiology, the distribution of sensory signals within the cord may be broader and less segment-specific than dermatomal maps suggest, leaving the segment-level localization of sensory-evoked activity in humans uncertain. Spinal cord functional magnetic resonance imaging (fMRI) is currently the only technique capable of noninvasively mapping sensory activity with high spatial resolution in the human spinal cord. However, its application remains technically challenging and is limited by the uncertainty in segmental localization.

In this study, we leveraged recent advancements in spinal cord fMRI, including spinal nerve rootlet–based spatial normalization, to investigate how sensory information is represented and distributed within the human spinal cord during electrocutaneous stimulation of the third digit of the right hand (i.e., C7 dermatome). Forty healthy adults were scanned with electrocutaneous stimulation at four individualized intensities across multiple runs to quantify (i) the rostrocaudal distribution of sensory-evoked activity, (ii) intensity-dependent changes in detectability and localization, and (iii) the effect of normalization strategy on segmental localization.

Across participants, stimulation produced activation localized in the lower cervical cord (e.g., C6-C8), with the most consistent segmental localization near C7. Stronger stimulation increased detectability and produced more consistent segmental localization across participants. Importantly, normalization that incorporated nerve rootlet landmarks sharpened localization and improved sensitivity relative to conventional intervertebral disc–based alignment. This highlights the value of functionally relevant anatomical landmarks for group inference in the spinal cord. Responses were strongest in the initial run and attenuated with repetition, suggesting habituation or adaptation that can bias multi-run paradigms if unmodeled. Together, our results define practical acquisition and analysis conditions (e.g., stimulation strength, anatomical alignment strategy, and run structure) under which segment-level spinal sensory responses can be detected, thereby supporting more reliable studies of human spinal cord future basic and translational studies, including pain mechanisms, sensory function, and spinal injury.

## 1. Introduction

Sensory organization at the spinal segment level is commonly inferred using dermatomal maps that assume a fixed correspondence between cutaneous regions and spinal segments. Conventional dermatomal maps, largely derived from manual drawings, vary considerably and fail to account for inter-individual variability (Lee et al., 2008). Moreover, there remains ambiguity regarding what dermatomes actually represent anatomically—spinal nerves, spinal cord segments, dorsal root ganglia, or dorsal horns (Lee et al., 2008).

Modern spinal anatomy and physiology suggest that sensory-evoked activity may be broader and less segment-specific than dermatomal maps imply. Afferents entering a given dorsal root can arise from multiple peripheral nerves, and single nerves can contribute to multiple roots; moreover, intersegmental anastomoses between cervical dorsal roots (present in ∼61% of individuals) can shift entry to adjacent segments (Lissarrague et al., 2025; Moriishi et al., 1989). After entry, primary afferents may ascend or descend several segments within Lissauer’s tract before terminating in the dorsal horn, further widening the rostrocaudal spread of input (Coggeshall et al., 1981; Light & Perl, 1979; Sugiura et al., 1986). Within the spinal cord, sensory signals are transformed by interneuronal spinal networks (Chung & Coggeshall, 1983; Nathan & Smith, 1959; Pierrot-Deseilligny & Marchand-Pauvert, 2002; Soteropoulos et al., 2013; Willis & Westlund, 1997) and descending supraspinal pathways (Stenner et al., 2025) that relay, integrate, and modulate input within and between spinal segments. The spatial location of sensory activity may also depend on the stimulus intensity. In the spinal cord, it remains unclear whether intensity is reflected primarily by increased firing rates, recruitment of additional afferent fibers, engagement of broader interneuronal and supraspinal pathways, or some combination of these mechanisms, particularly across the transition from innocuous to noxious sensation (Summers et al., 2010).

Together, these considerations raise fundamental questions regarding the spinal cord’s role in sensory processing and the validity and reliability of dermatomal maps used clinically: How spatially localized are sensory representations in the human spinal cord, how does their extent change with stimulus intensity, and how much of the observed pattern reflects segmental organization versus distributed spinal processing? Addressing these questions requires methods capable of measuring spinal cord activity in humans with sufficient spatial specificity.

Spinal cord functional magnetic resonance imaging (fMRI) is the only technique to noninvasively map sensory activity with high spatial resolution in the human spinal cord. Using blood-oxygen-level–dependent (BOLD) signals as a proxy for neural activity, spinal cord fMRI can be used to localize and quantify the spatial extent of spinal activity, but remains technically challenging (Kinany et al., 2022). A persistent methodological challenge has been spatial normalization, which is a method used to align anatomy across individuals and an essential step for group-level analyses. Spinal cord fMRI studies have relied on spine anatomy (i.e., vertebral or intervertebral disc levels) for spatial normalization. This approach is inherently inaccurate because of varying correspondence between vertebral and spinal cord segment levels across individuals due to differences in spinal cord length relative to the vertebral column (Cadotte et al., 2015). To overcome this challenge, we recently introduced a spinal nerve rootlet–based spatial normalization method, designed to improve the alignment across individuals by directly targeting neuroanatomical landmarks relevant to segmental organization. This approach improved group-level localization of motor task-related fMRI activity but has not yet been evaluated using a spatially localized sensory stimulus (Bédard, Valošek, et al., 2025).

In this study, we leverage recent advancements in spinal cord fMRI including rootlet-based spatial normalization to investigate how sensory information is represented and distributed within the human spinal cord during electrocutaneous sensory stimulation of the third digit of the right hand (i.e., C7 dermatome) at multiple individualized intensities across multiple stimulation runs. We first evaluate the impact of anatomical alignment based on spinal segment levels. We then characterize intensity-dependent encoding, and finally, we assess response reliability and the presence of spinal habituation or sensitization. Together, these experiments test how anatomical alignment, stimulus intensity, and repeated exposures shape the apparent segmental organization, spatial extent, and reliability of sensory-evoked spinal activity, providing new insight into human sensory processing in the spinal cord. These results also define practical acquisition and analysis conditions for more reliable segment-level mapping of human spinal sensory processing.

## 2. Materials and methods

### 2.1. Participants

Forty healthy adult volunteers were recruited (age = 38.8 ± 14.52, 50% female). Exclusion criteria included contraindications to MRI, pain conditions, opioid use, previous spine surgery, ongoing spine conditions or known peripheral nerve compression, major physical trauma to the spine, cancer, and major medical, psychiatric, and neurological conditions. Inclusion criteria included an age between 20 and 79 years old, right-handedness, and fluency in English. Eligible volunteers were informed about the study protocol and provided written informed consent before participation. All participants received compensation, and the study was approved by the Stanford University Institutional Review Board.

### 2.2. Image acquisition

All imaging was performed in a supine position on a 3T GE SIGNA Premier scanner equipped with a custom 56-channel head-neck coil (designed and built by NeuroPoly Lab, Polytechnique Montreal) at the Richard M. Lucas Center for Imaging at Stanford University. A SatPad cervical collar was also used to improve B0 field homogeneity (De Leener et al., 2020). Physiological measurements were collected using a pulse oximeter placed on the left index finger and a respiratory belt positioned around the abdomen. Spinal cord fMRI BOLD data was acquired using a 2D (axial) spatially selective reduced field-of-view (FOV) pulse GE-EPI sequence with dynamic per-slice shimming (21 slices, 1.25 mm × 1.25 mm × 4.00 mm, TR = 1.7 s, bandwidth = 250 kHz, TE = 30 ms) (Islam et al., 2019). The pulse sequence was designed and implemented under the KSFoundation^18^. The FOV was centered at the C5-C6 intervertebral disc and oriented obliquely to remain perpendicular to the spinal cord, spanning approximately from C5 to T1 spinal segment levels.

For shimming, both axial (FOV = 22 cm, matrix size = 128 × 128, ΔTE = 1 ms) and sagittal (FOV = 30 cm, matrix size = 256 × 64, ΔTE = 1 ms, one slice, during a 3 s breath-hold) B0 field maps were acquired to calculate slicewise shims using the methods proposed by Islam et al. (2019). A high-resolution sagittal T2-weighted (T2w) structural scan was acquired for spatial normalization using a 3D turbo spin-echo sequence (CUBE) with the following parameters: TR = 2.5 s, TE = 128 ms, matrix size: 64 × 320 × 320, resolution: 0.62 mm × 0.62 mm × 0.62 mm, bandwidth = 520.8 kHz) (Cohen-Adad et al., 2021). This image enables clear visualization of the spinal rootlets (Valošek et al., 2024) and intervertebral discs.

### 2.3. Functional MRI experiment

An overview of the experimental setup is provided in **Figure 1**. Electrocutaneous stimulations were applied to the third digit of the right hand (C7 dermatome) using pre-gelled Ag/AgCl ring electrodes with 50 cm lead wires (Cadwell Industries, Kennewick, WA, USA) and MRI-compatible Biopac system. Four individualized stimulation intensities, evenly spaced between each participant’s sensory threshold and maximum tolerable intensity, were used for the fMRI experiment. Thresholds were determined in a single ascending series (0.2–10 mA, 0.2 mA steps) using a 2 s on / 2 s off pattern. Participants squeezed a hand-held ball at first sensation (sensory threshold) and again at their maximum tolerable intensity; pain ratings were collected at both points. Subsequent stimulation levels were selected between 0.2 mA above detection and 0.2 mA below tolerance.

**Figure 1.**
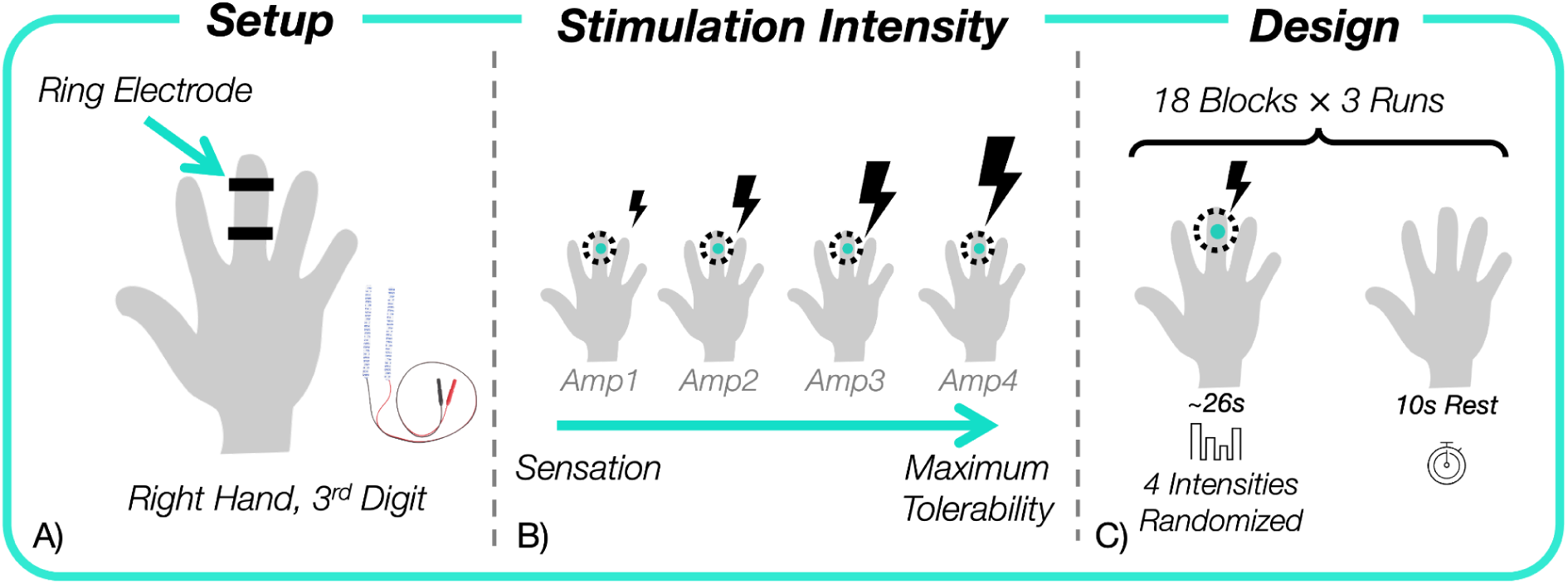
Experimental setup. **A)** MRI-compatible Biopac system and Ag/AgCl ring electrodes on the third digit of the right hand. **B)** Four individually determined electrocutaneous stimulation intensities between sensory threshold and maximum tolerable intensity. **C)** Each run (∼10 min, 376 volumes) included 18 blocks, with 10 seconds rest periods. Each block delivered four randomized monophasic pulses (100 Hz, pulse width = 0.2 ms, duration = 5 s).

Each run consisted of 18 stimulation blocks, each separated by a 10-second rest period. Within each block, four stimulations were delivered using monophasic pulses (100 Hz, pulse width = 0.2 ms, duration = 5 s), with the order of the intensity levels randomized within the block and interleaved by 2-second intervals. Participants completed three runs, each lasting 10 minutes, for a total of 376 fMRI volumes. After each run, participants reported perceived pain intensity on a verbal 0–10 scale with anchors of no pain (0) and worst pain imaginable (10).

### 2.4. Data processing

#### 2.4.1. Preprocessing

An overview of the processing pipeline is presented in **Figure 2** and available on Github: https://github.com/kennethaweberii/Dermatomal_Mapping_R01/releases/tag/r20260213 The pipeline includes motion correction, physiological noise removal, slice-timing correction, temporal filtering, and spatial normalization. The same preprocessing was applied to all three runs and detailed below.

**Figure 2.**
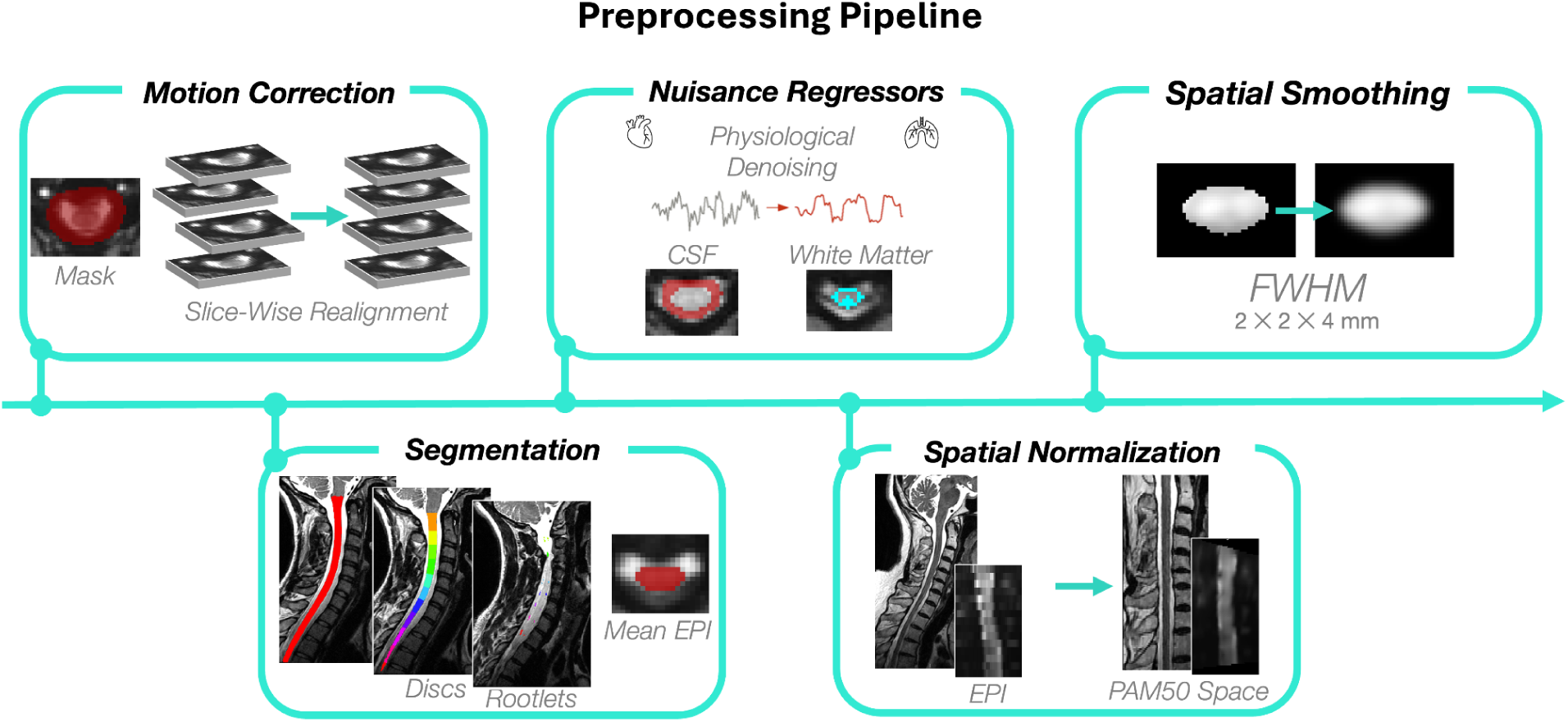
Processing pipeline for fMRI of the spinal cord. Slicewise motion correction of the EPI time series was applied within a mask surrounding the spinal cord. The spinal cord, nerve rootlets, and intervertebral discs were segmented/labeled from the T2w image and the spinal cord was segmented from the mean motion corrected EPI image. Physiological noise, cerebrospinal fluid, and white matter signals were removed. The T2w image was registered to the PAM50 template based on rootlets and discs, and spatial smoothing was applied to the EPI image in the PAM50 template space.

Image processing was performed using the Spinal Cord Toolbox (SCT) v7.0 (De Leener et al., 2017) and FSL v6.0 (Jenkinson et al., 2012). The functional spinal cord time series was first motion corrected using FSL’s `FLIRT` tool (Jenkinson et al., 2012) in two steps. A mask around the spinal canal was created by dilating the automatic segmentation of the spinal cord obtained using `sct_deepseg` `sc_epi` (Banerjee et al., 2025), to restrict the analysis to the spinal column. First, each axial slice of the functional time series was realigned to the mid-volume using 2D rigid-body registration. A mean image was computed, and all volumes were realigned to this mean image in a second round of motion correction (Weber et al., 2014). Each step was visually inspected to ensure accurate alignment across volumes using `sct_qc`. Motion outliers were identified using FSL’s `fsl_motion_outliers` and included as confound regressors in the first-level statistical analysis later. Mean temporal signal-to-noise ratio (tSNR) maps were computed following motion correction.

The spinal cord was segmented using `sct_deepseg` `sc_epi` on the motion-corrected mean functional image (Banerjee et al., 2025). The segmentation was visually assessed using `sct_qc` and manually corrected in cases of major errors (e.g.: under or over-segmentation) (Valošek & Cohen-Adad, 2024). A spinal canal mask was generated by combining the spinal cord and cerebrospinal fluid (CSF) segmentations from `sct_propseg` (De Leener et al., 2014). The CSF mask was computed by subtracting the corrected spinal cord mask from the canal mask.

Spatial normalization relies on the high-resolution T2w structural image. The spinal cord and cervical spinal rootlets (dorsal and ventral, from C2-T1 spinal segment levels) were automatically segmented using `sct_deepseg` `spinalcord` (Bédard, Karthik, et al., 2025; Karthik et al., 2025), and `sct_deepseg` `rootlets_t2`, respectively (Krejci et al., 2025; Valošek et al., 2024). Vertebral levels were also automatically labeled using `sct_label_vertebra` `to` allow further comparison of disc-based vs. rootlet-based normalization in fMRI analysis (Bédard, Valošek, et al., 2025). The spinal cord was registered to the PAM50 template using both disc- and rootlets-based approaches (`sct_register_to_template` `-lrootlet`) (Bédard, Valošek, et al., 2025; De Leener et al., 2018). **Figure 3** shows the correspondence between spinal and vertebral segments along with the registration pipeline. The resulting warping field served as an initial transform to align the PAM50 template to the mean functional image on a slicewise basis, using the spinal cord segmentation from the functional image. A white matter mask was derived by thresholding the PAM50 white matter probabilistic map at 0.9 in the native space.

**Figure 3.**
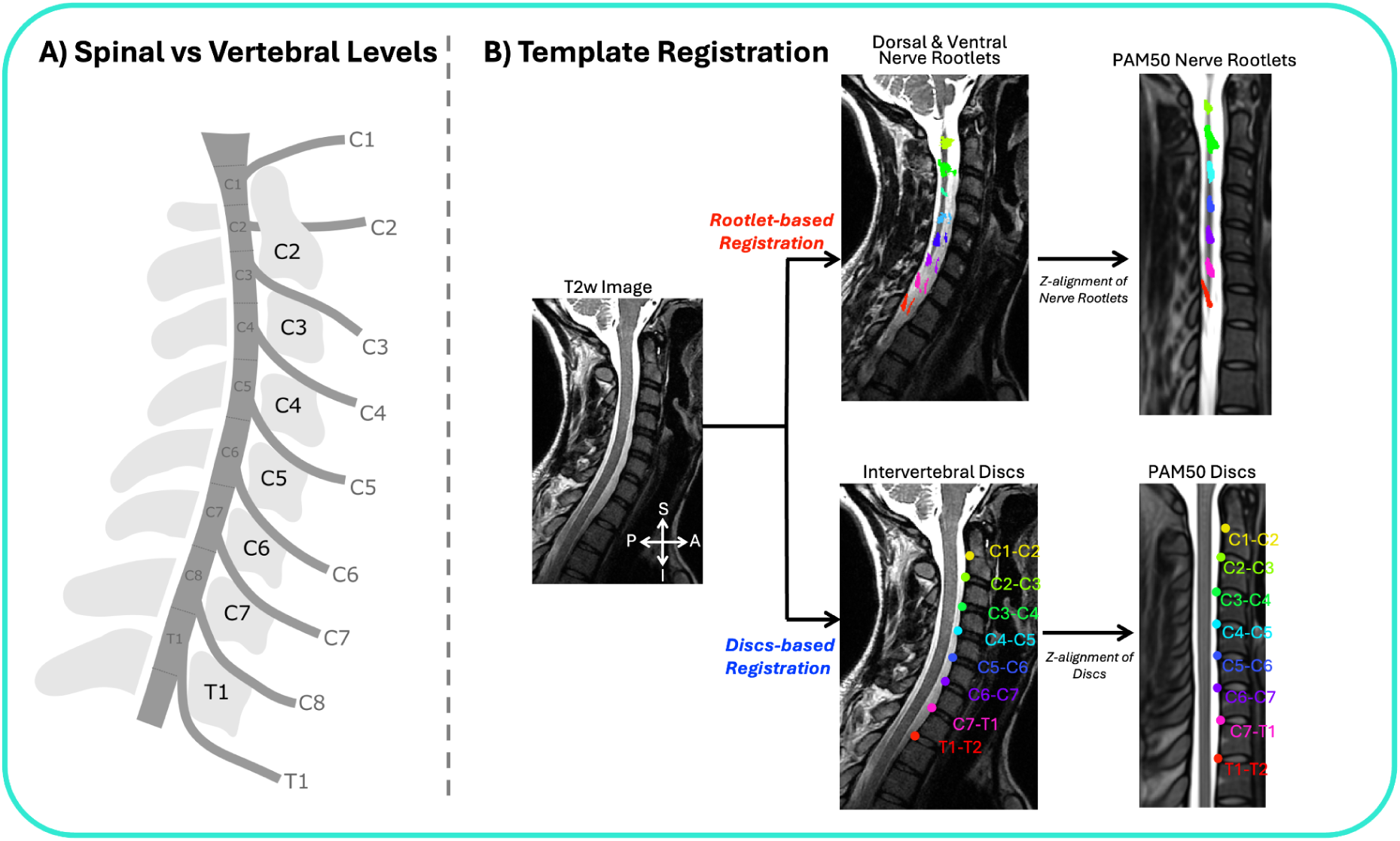
Template registration pipeline. **A)** Correspondence between spinal segment levels, nerve roots and vertebral level. **B)** Registration to the PAM50 template based on nerve rootlets (red) and based on intervertebral discs (blue).

To remove physiological and motion-related noise, slicewise nuisance regressors were generated. For CSF and white matter, the first principal components of their respective signal time series were extracted using `fslmeants`. Cardiac and respiratory signals were modeled using 32 regressors derived from PNM (Brooks et al., 2008), while motion-related fluctuations were accounted for by including slicewise x-axis and y-axis translations and z-axis rotations. All nuisance regressors (n = 37) were removed using a general linear model (GLM) implemented in FSL’s `FEAT`. Slice-timing correction was applied after denoising to account for inter-slice acquisition delays.

The resulting cleaned functional time series were warped to the PAM50 template space. In this template space (straightened cord), a Gaussian smoothing kernel (FWHM: 2 mm × 2 mm × 4 mm) was applied to primarily smooth along the rostrocaudal axis of the spinal cord, an effect that would be compromised in native space due to cord curvature. Additionally, high-pass temporal filtering with a 150-second cut-off was performed. This preprocessing pipeline yields standardized, denoised functional images ready for participant-level statistical analysis.

#### 2.4.2. Statistical inference

The preprocessed fMRI time series were analyzed using a single hemodynamic response function convolved with trialwise boxcar regressors corresponding to each stimulation intensity. This generated first-level activity maps for each run. For each participant, an average activity map across the three runs was computed using a second-level fixed-effects analysis (Jenkinson et al., 2012). Individual participant maps were then entered into the group-level analyses using nonparametric permutation testing with FSL’s randomise, with a cluster-level, family-wise error (FWE) correction (Z score > 2.30, FWE p < 0.050) (Winkler et al., 2014). Age and sex were included in the group-level analyses as covariates of no interest. Analyses were restricted to the region of the spinal cord covered by each participant’s functional acquisition FOVs, which spanned approximately from C5 to T1 spinal cord levels.

Each stimulation intensity level was investigated separately, and a linear contrast across the four stimulation intensities was modeled at the group level to examine how stimulus strength modulates BOLD responses. Laterality was assessed with the left-right ratio (LR) defined by:

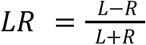

With LR defined as the left-right ratio, L, the number of active voxels in the left hemicord, and R, the number of active voxels in the right hemicord.

#### 2.4.3. Region of interest analysis

Region-of-interest (ROI) analyses were performed within the spinal cord gray matter (GM). The GM mask was generated by binarizing the PAM50 GM atlas at a 0.5 threshold. Right and left hemicord ROIs were created by summing the corresponding PAM50 atlas regions (https://spinalcordtoolbox.com/stable/user_section/tutorials/atlas-based-analysis/adding-custom-tracts.html). ROIs were generated for the right GM, left GM, and all the spinal coverage of the group functional scans, within spinal segment levels C6, C7 and C8.

For each participant, the number of active voxels were extracted and mean Z scores were computed within each ROI for all four stimulation intensities. ROI-level statistical analyses included paired t-tests to compare activity between hemicords and linear models to assess changes across stimulation intensity.

#### 2.4.4. Test-retest reliability

Test-retest reliability was assessed using intra-class correlation (ICC) to quantify the stability of fMRI responses across the three runs. Voxelwise ICC was computed using a two-way mixed-effects model, and ICC(3,k) was used to evaluate the reliability of the average activity across the three runs, following Caceres et al. (2009). ICC values were interpreted according to Cicchetti’s (1994) guidelines (<0.40 poor, 0.40-0.59 fair, 0.60-0.74 good, 0.75-1.00 excellent). To further assess replicability, the dataset was split by run and participant- and group-level activity were analyzed. This analysis was done only on the highest stimulation amplitude (Amp4).

#### 2.4.5. Sample size analysis

We performed a Monte Carlo-based sample size analysis to assess the robustness of group-level activity patterns. Group analyses with varying sample sizes were performed on the average of the three runs with the Amp4 stimulation intensity, using FSL *randomise* with a voxelwise threshold of Z > 2.3 and FWE cluster correction (p < 0.050). Subsamples of size 10, 15, 20, 25, 30, 35, and 40 were drawn without replacement, and each subsample analysis was repeated for 1,000 iterations.

## 3. Results

TSNR was similar across the three runs and the average of the runs, but was generally lower at the most inferior spinal segment level (T1) (**Figure 4**).

**Figure 4.**
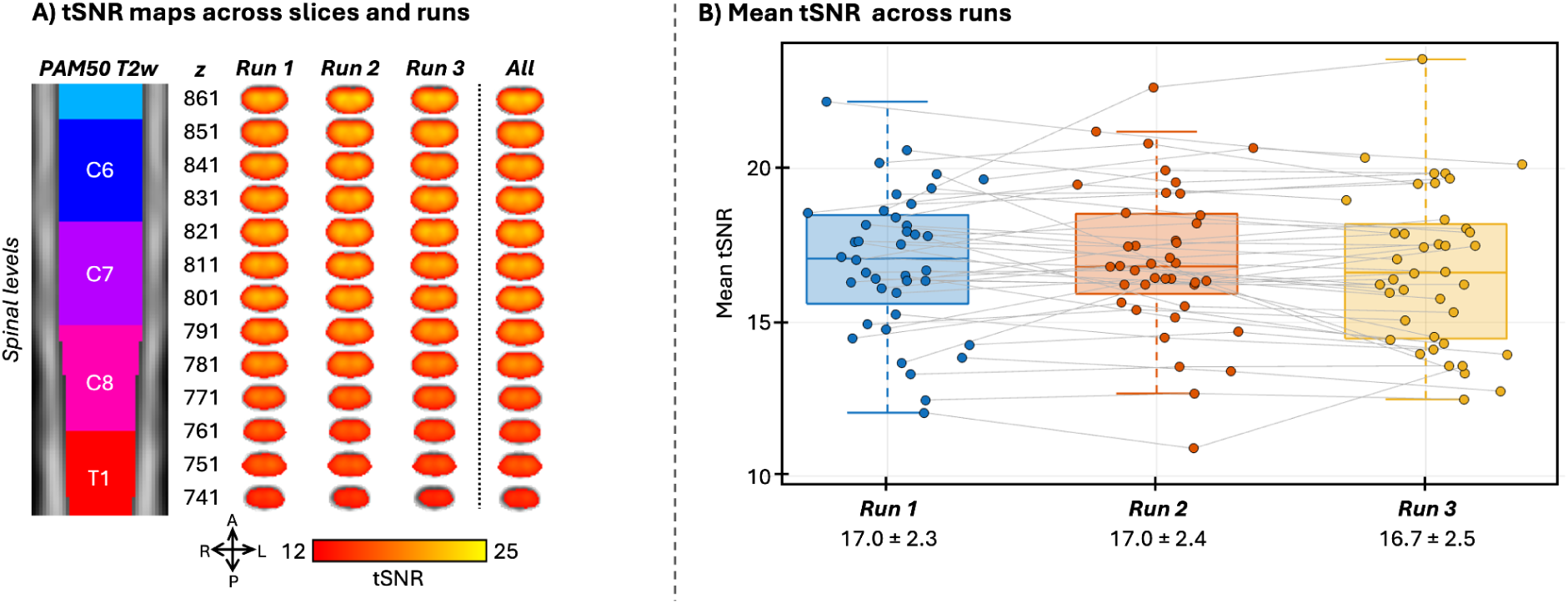
Temporal signal-to-noise ratio (tSNR) after motion correction. **A)** tSNR map in the PAM50 template space for each run and across all runs after motion correction. The mean column represents the average tSNR within the spinal cord mask, across all runs. Rootlets-based spatial normalization was used to generate the group level tSNR map. tSNR was computed in the native space, after motion correction (linear interpolation) for each run, and warped to the template with linear interpolation. Group tSNR is similar across the 3 runs, while it decreases at the inferior field-of-view. **B)** Mean tSNR across runs after motion correction in te native space. Each dot represents one participant. Mean tSNR and standard deviation are reported under each run. No significant differences were found across runs (Bonferroni-corrected paired t-test)

### 3.1. Impact of anatomical landmark for registration

We investigated the impact of anatomical landmarks on functional group activity maps using either spinal nerve rootlets or intervertebral discs labels during template registration (see **Figure 3**). A mean T2w image (N = 40) registered to the template shows sharper delineation of the rootlets when using rootlets-based compared to disc-based registration (see **Figure S1**). For the functional results, group-level maps for Amp4 comparing both rootlet and discs-based registration are presented in **Figure 5**. Rootlets-based registration increased Z scores and the number of active voxels, yielding more spatially localized clusters at C7 compared to disc-based registration. The same improvement was found for the linear contrast (see **Figure S2**).

**Figure 5.**
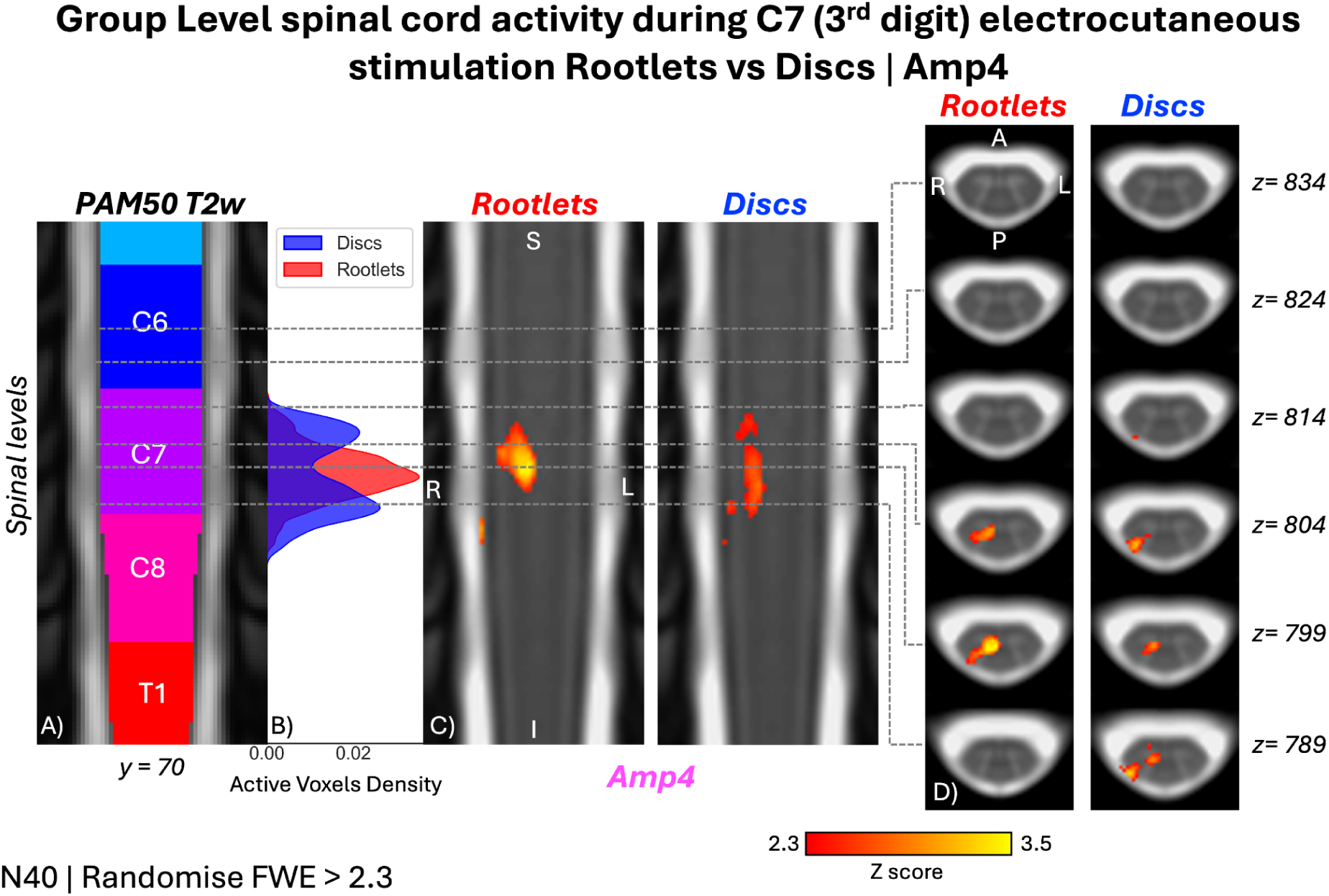
Group-level activity maps during electrocutaneous stimulation of the 3rd digit of the right hand (N = 40, Amp4) comparing rootlets- (red) vs. discs-based (blue) registration. **A)** Coronal view (y = 70, PAM50) with spinal segment levels. **B)** Active voxel density plot. **C)** Coronal slices with activity overlay. **D)** Axial views of representative clusters. Maps: Randomise, Z score > 2.3, and cluster corrected for family-wise error (FWE) at p < 0.050. In the coronal view, clusters appear narrower with a higher Z score, whereas the disc is more spread out.

### 3.2. Stimulation intensity

For the lowest stimulation intensity (Amp1), with an average amplitude of 2.4 ± 0.9 mA, the reported pain intensity was 0.2 ± 0.5. For the highest stimulation amplitude of 6.5 ± 3.0 mA (Amp4), the mean reported pain intensity was 4.41 ± 2.75.

#### 3.2.1. Intensity encoding at the participant level

At the participant level, the BOLD signal increased significantly with stimulation intensity in the right dorsal horn, particularly at C6–C7 spinal segment levels, as reflected by greater number of active voxels and higher average Z scores across intensities, and significant linear fit (**Figure 6**). No significant increase was found in the number of active voxels or in the average Z score across intensities in the left GM.

**Figure 6.**
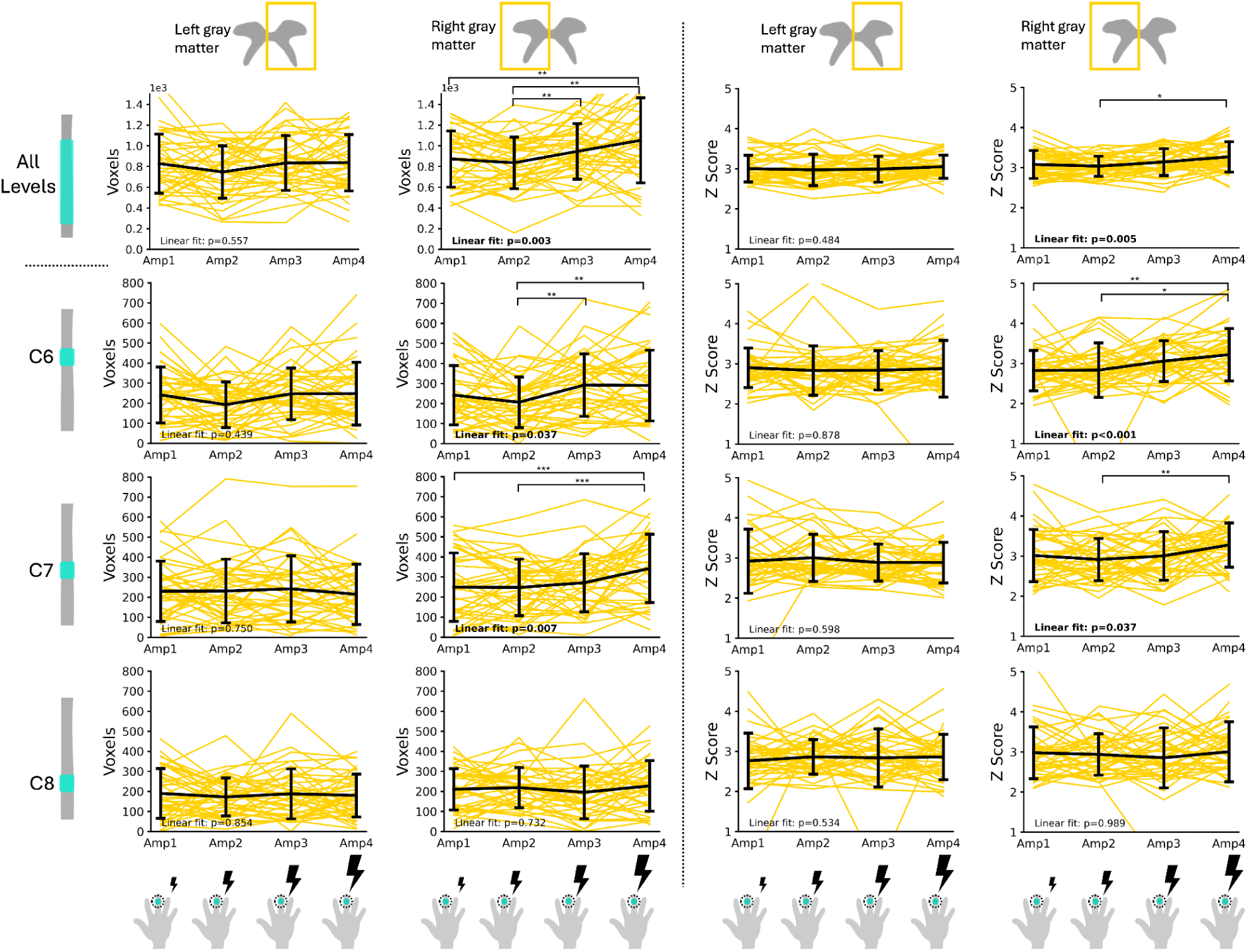
Participant-level active voxels and average Z score across stimulation intensities. Number of active voxels and average Z score within left and right spinal cord gray matter across stimulation intensities for all spinal segment levels, and specifically C6, C7, and C8 (p < 0.050). Yellow lines represent one individual participant; the black line shows the group average, with brackets indicating standard deviations. Significance: *p < 0.050, **p < 0.010, ***p < 0.001 (Bonferroni-corrected paired t-test). A significant linear increase of active voxels and average Z score is observed in the right gray matter across all spinal segment levels, and specifically at C6 and C7.

#### 3.2.2. Intensity encoding at the group level

All group level results are shown using rootlets-based registration methods, or mentioned explicitly if otherwise.

At the group level, the maximum intensity (Amp4) showed significant clusters of activity localized to the right dorsal horn at C7 spinal segment level (Z score > 2.3; FWE p < 0.050). Intensities Amp1, Amp2 and Amp3 did not yield any significant clusters that passed FWE correction. Uncorrected maps for each stimulation intensity are provided in the supplementary material (**Figure S3**). In addition, the BOLD signal increased linearly with stimulation intensity, localized to the caudal end of C6 and C7 spinal segment levels and the right dorsal horn (i.e., negative LR ratio) (**Figure 7**). No relationship between sex, age and spinal activity were found.

**Figure 7.**
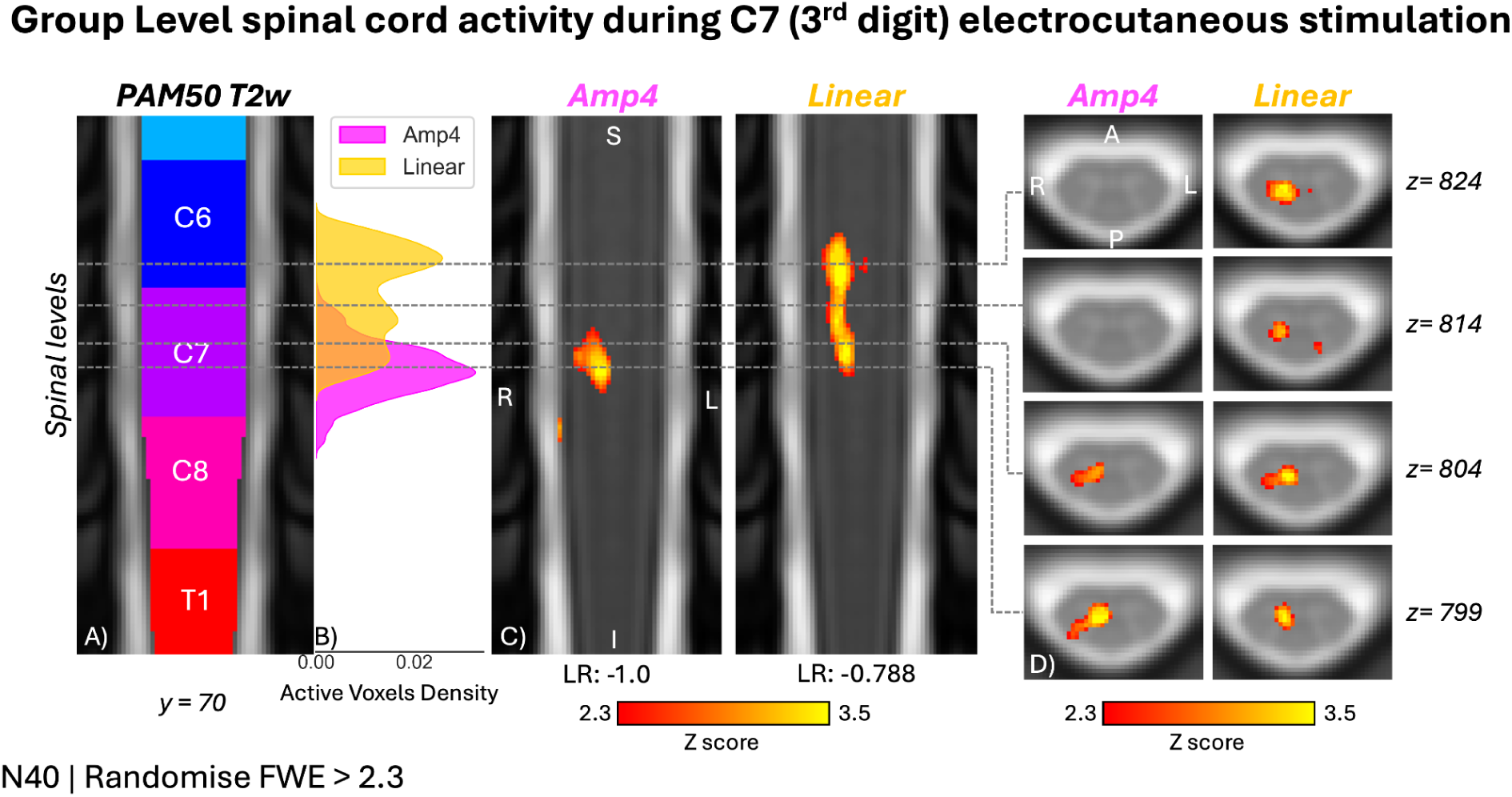
Group-level activity maps during electrocutaneous stimulation of the 3rd digit of the right hand (N = 40) with various stimulation intensities. **A)** Coronal view (y = 70 on the PAM50 referential system) with an overlay of the spinal segment levels, matching the coverage of the functional EPI scans. **B)** Active voxel density for stimulation intensity Amp4 and linear fit between stimulation intensities. **C)** Coronal slices with an overlay of the activity maps. The location of the activity was assessed using the left-right (LR) ratio. **D)** Axial views of representative activity clusters. The activity maps were generated using a voxel-wise thresholded at Z score > 2.3, and cluster corrected for family-wise error (FWE) at p < 0.050. Simulation amplitudes Amp1, Amp2 and Amp3 did not yield any significant clusters that passed FWE correction, and are not shown here; uncorrected maps for each stimulation intensity are provided in the supplementary material (**Figure S3**). Rootlets-based spatial normalization was used to generate the group level maps.

### 3.3. Test-retest reliability

As stimulation intensities Amp1, Amp2 and Amp3 did not yield any significant clusters at the group level, we mainly focus on stimulation intensity Amp4 for the subsequent analysis to investigate the test-retest reliability across runs. All group level results are shown using rootlets-based registration methods, or mentioned explicitly if otherwise.

#### 3.3.1. Intra-class-correlation across runs

We first report voxelwise ICC across the three runs and participants for stimulation intensity Amp4 (**Figure 8**). ICC was generally low, but showed a higher ICC localized at C7 spinal segment level, in the right dorsal horn around 0.5, which corresponds to fair reliability (Cicchetti, 1994).

**Figure 8.**
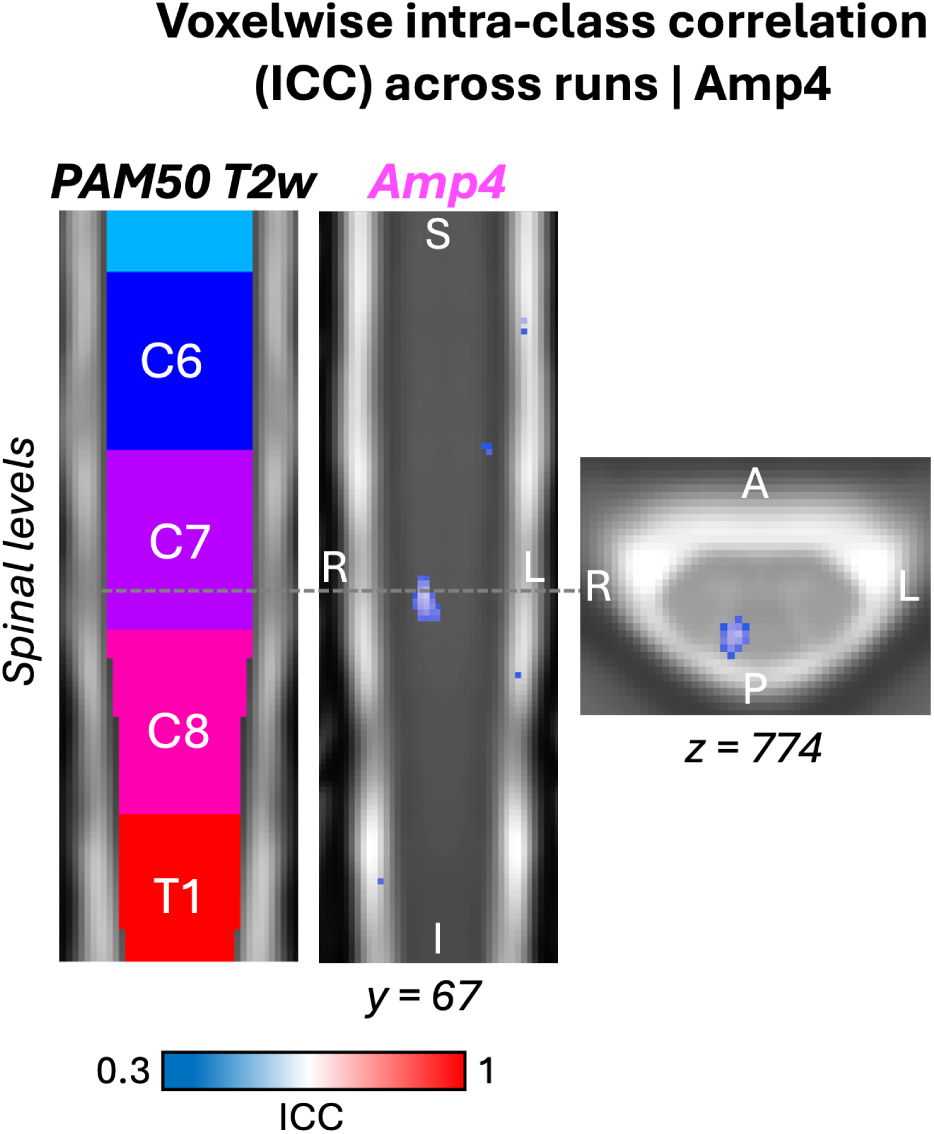
Voxelwise intra-class correlation (ICC) across the three runs and participants for stimulation intensity Amp4. The PAM50 template with overlaid spinal segment levels is shown on the left. A coronal slice (y = 67) with the corresponding ICC map is displayed in the center, and an axial slice (z = 774) with ICC overlay is shown on the right. Higher ICC values indicate greater consistency of activity across runs. Group-level ICC maps were generated using rootlet-based spatial normalization. ICC(3,k) was computed quantifying the consistency of activity across the three runs for each participant.

#### 3.3.2. Participant level analysis across runs

To investigate the low reliability across runs, we examined Amp4 responses at the participant level by quantifying the number of active voxel and average Z score within the GM across spinal segment levels for each of the three runs. As shown in **Figure 9**, the number of active voxels decreased linearly across runs at all segment levels, whereas average Z scores did not change significantly. To determine whether this decrease of active voxels emerged within a single run, we also assessed active voxel counts and average Z scores across trials in run-1 (see **Figure S4**) and found no significant within-run trend. Potential confounds were evaluated and did not support alternative explanations: motion outliers did not increase across runs (**Figure S5**), and maximum reported pain intensity was stable across runs (**Figure S6**).

**Figure 9.**
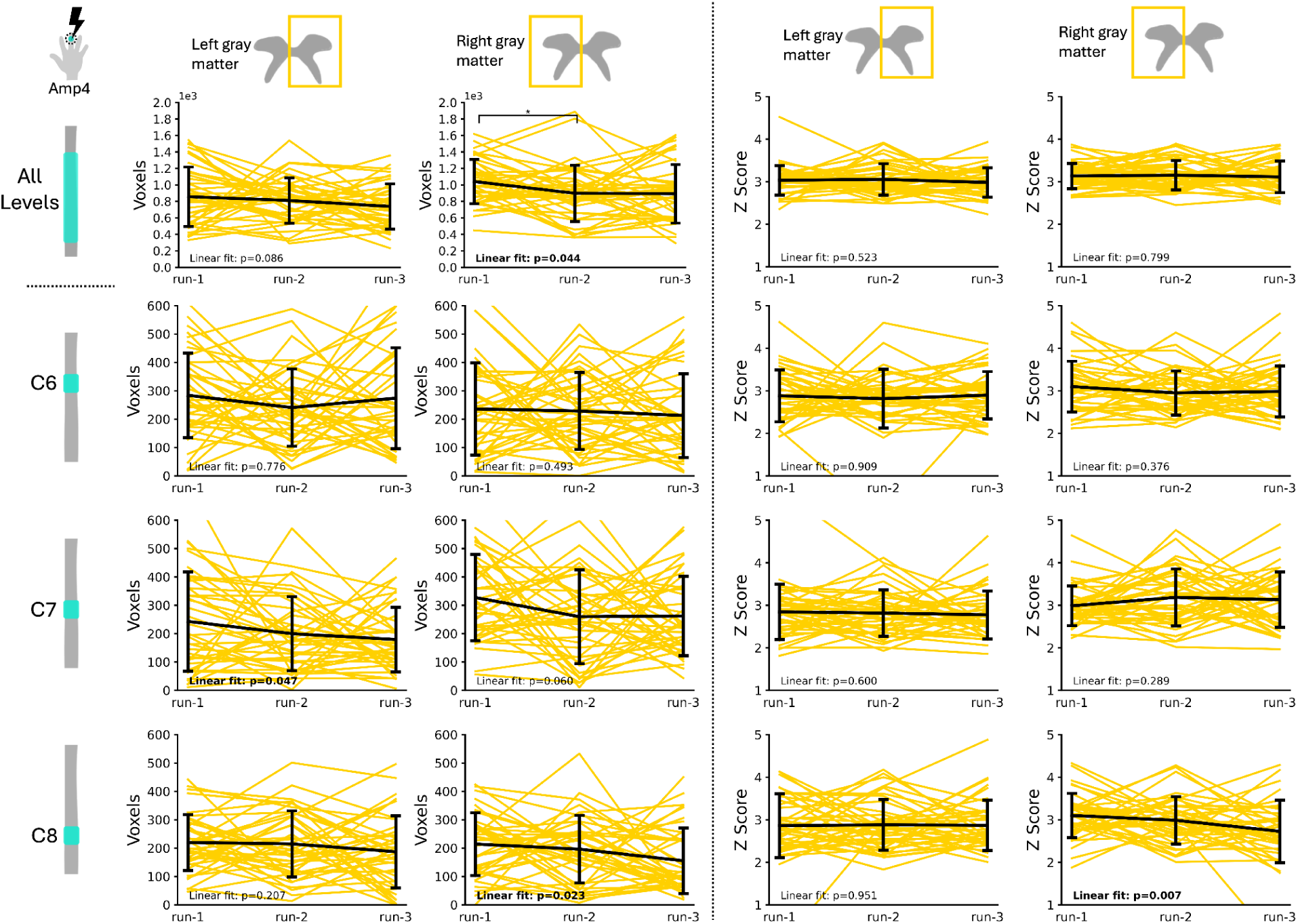
Average number of active voxels and average Z score within left and right spinal cord gray matter across runs for Amp4 and all spinal segment levels, C6, C7 and C8. Each yellow line represents one participant as the black line represents the average across all participants with the brackets showing the standard deviation. Bonferonni-corrected paired t-test. *: p < 0.050; **: p < 0.010; ***: p < 0.001. The number of active voxels decreases between runs across all levels and at C7 spinal segment level. The average Z score did not change significantly across runs.

#### 3.3.3. Group level analysis across runs

We next examined group-level activity separately for each run. Run-1 showed significant clusters for Amp3, Amp4 and linear contrast (**Figure S7**; randomise, Z score 2.3, FWE-corrected p < 0.050), matching the pattern obtained when averaging across all three runs (see **Figure S8**). In contrast, runs 2 and 3 did not produce any FWE-corrected significant clusters for any intensity. Uncorrected maps (**Figure 10**) nevertheless suggest a reduction in the Amp4 response after run-1, with fewer active voxels and lower Z score in runs 2 and 3, consistent with the density distributions of active-voxel.

**Figure 10.**
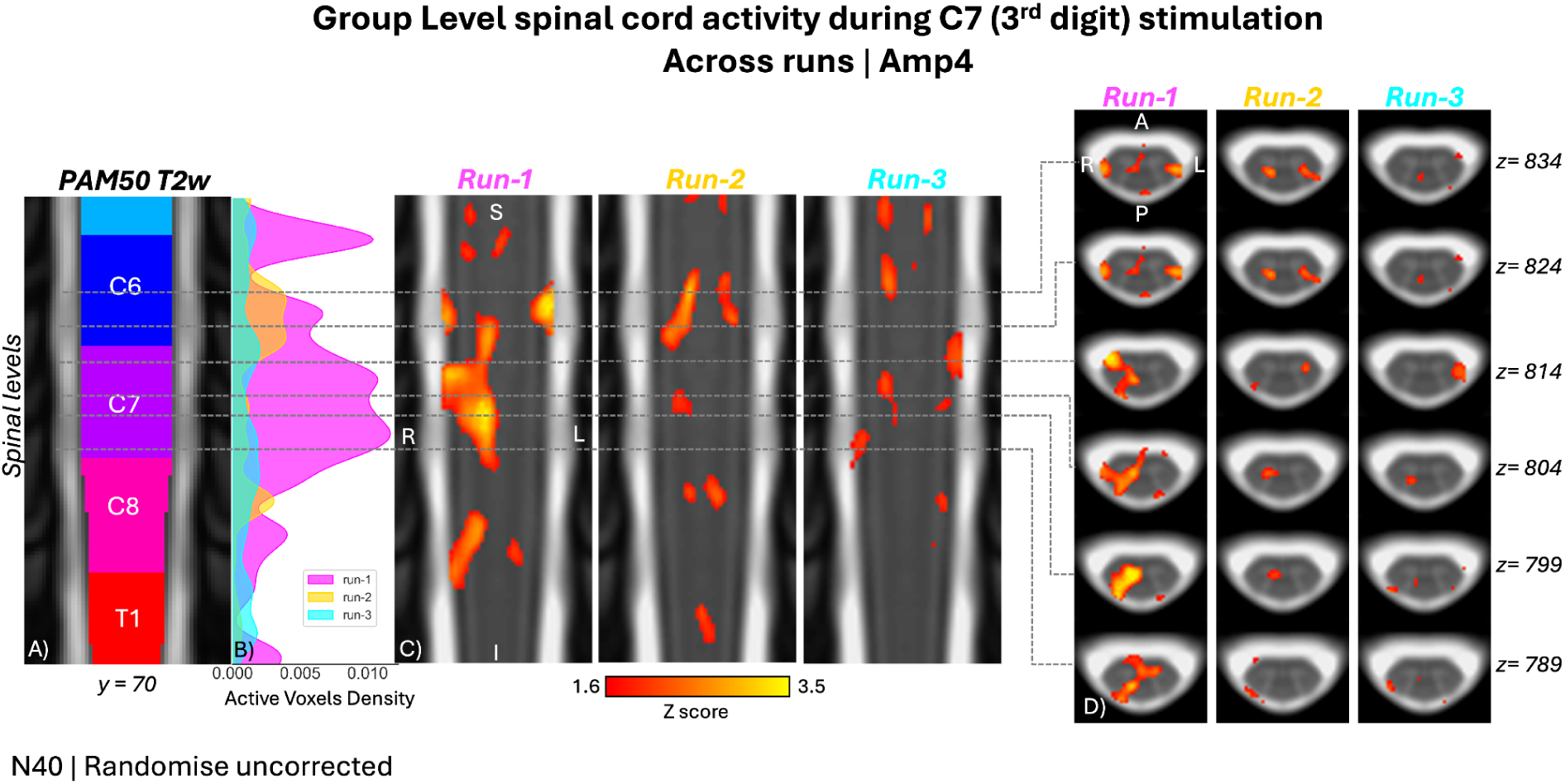
Group-level activity maps during electrocutaneous stimulation of the 3rd digit of the right hand (N = 40) with Amp4 across individual runs. **A)** Coronal view (y=70 on the PAM50 referential system) with an overlay of the spinal segment levels, matching the coverage of the functional EPI scans. **B)** Density plot of active voxels for stimulation amplitude Amp4 across three runs. **C)** Coronal slices with an overlay of the activity maps. **D)** Selected axial views centered on representative activity blobs. The activity maps were generated using a Randomise model, voxel-wise thresholded at Z score > 1.6, and uncorrected for family-wise error (FWE). Rootlets-based spatial normalization was used to generate the group level maps. A lower density of active voxels is observed between run-1 vs run-2 and run-3.

### 3.4. Sample size

The Monte Carlo analysis provided an empirical estimate of the sample size required to reliably detect segment-specific activity (Amp4, across all runs). We observed that the activity cluster at C7 spinal segment level was detected above 50% of the time with a sample size of N = 28 (**Figure S9**). By repeatedly resampling the dataset across increasing group sizes, we observed a clear improvement in detection frequency with larger cohorts. It is important to note that, since there were 1000 iterations, there were some repetitions of sample for higher sample size.

## 4. Discussion

Our findings identify practical conditions under which segment-level spinal sensory responses can be detected with spinal cord fMRI. Across participants, electrocutaneous stimulation of the third digit of the right hand was most consistently localized near the C7 spinal cord segment. Detectability increased with stimulation intensity. Critically, group inference was improved by anatomical normalization that incorporated rootlet landmarks, supporting the view that functionally relevant anchors can reduce artifactual spread introduced by disc-based alignment. At the same time, responses attenuated across repeated runs, indicating that repetition can materially alter measured effects and should be explicitly modeled or mitigated in the study design. Together, these results help reconcile prior reports of both focal and distributed activation and provide a framework for more reliable spinal fMRI studies of human sensory processing.

### 4.1. Impact of anatomical landmark: disc vs. nerve rootlet reference

An important step in mapping dermatomes with fMRI is the choice of anatomical landmarks used during spatial normalization (Kinany et al., 2022). Aligning participants by vertebral or disc levels can decrease the reliability of group-level maps because levels based on spine anatomy are an indirect proxy for the anatomy of interest, the spinal segment levels. In contrast, normalizing data using spinal nerve rootlets better aligns corresponding spinal segments across participants and therefore improves functional correspondence, as reflected by increased number of active voxels, average Z-score and more localized clusters of activity in the group-level maps (**Figure 5**).

In previous work, we reported that rootlet-based registration improved alignment compared with disc-based registration in a task-based motor fMRI study (Bédard, Valošek, et al., 2025). However, motor tasks typically recruit multiple myotomes and span several spinal segments, producing spatially broad activation. As a result, it was difficult to determine whether the improved registration truly enhanced segment-level localization, since the “ground truth” activation was not expected to be focal. Here, using a localized sensory stimulus that engages a more restricted segmental representation, we can demonstrate a clearer improvement in group-level localization.

### 4.2. Mapping of dermatomes with electrocutaneous stimulation

#### 4.2.1. Spatial localization

At both the participant and group levels, we observed a linear increase of BOLD responses with higher stimulation intensities, primarily localized to the right GM at the C7 spinal segment level and extending partially to the caudal end of C6 (**Figure 6** and **Figure 7**). This localization is consistent with the expected dermatome (3rd digit stimulation, C7 dermatome). The activity extending to C6 spinal segment level could be associated with the fact that afferents entering via the C7 nerve root do not necessarily synapse at the C7 spinal cord segment; fibers can ascend or descend several levels within Lissauer’s tract before terminating in the dorsal horn (Coggeshall et al., 1981; Light & Perl, 1979; Sugiura et al., 1986). We did not observe a comparable intensity-dependent response at the C8 spinal segment level, which could be due to the lower tSNR measured at C8 (**Figure 4**), where proximity to the lungs increases field inhomogeneity, susceptibility artifacts, and signal loss (Finsterbusch, 2014).

In the present study, the use of rootlet-based spatial normalization likely improved segment-level correspondence across participants, which may have contributed to the more localized group-level activity observed here around C7 compared to previous studies. For example, Stracke et al. (2005) used electrical stimulation targeting the C6, C7, and C8 dermatomes at 1.5T and observed activity not only at corresponding levels but also at C3 and C4, which was attributed to the potential involvement of propriospinal interneurons. More generally, Landelle et al. (2021), noted in a review that spinal fMRI activity often peaks at the expected dermatome level but typically spans multiple segments. Consistent with this, Weber et al. (2020) applied tactile stimulation to the lateral shoulder (C5) and the dorsal surface of the third digit (C7), and observed activity extending across multiple segments in both conditions. Similarly, Lawrence et al (Lawrence et al., 2008) used vibration across several dermatomes (C5, C7, C8, T1, L4, S1) and found peak activity at expected levels alongside spread to adjacent segments and bilateral dorsal and ventral activity. Differences across studies may also reflect the stimulation site and intensity; however, because modalities vary (electrocutaneous, tactile, vibration), stimulus intensity is difficult to compare directly. Some of the apparent rostrocaudal spread reported in prior work may also reflect methodological factors, particularly the use of vertebra-based alignment as a proxy for spinal segments. Because vertebral levels do not map consistently onto spinal cord segments across individuals, imperfect segmental alignment at the group level can blur responses and produce artificially widespread activity.

Axially, both the highest stimulation intensity (Amp4) and linear-contrast clusters were localized to the anterior portion of the dorsal horn, consistent with expected sensory pathways. Aβ afferents terminate primarily within the intermediate dorsal horn laminae (i.e., laminae IV), which aligns with the observed activity pattern (**Figure 7**). However, the limited axial resolution and the use of in-plane spatial smoothing likely restricted our ability to resolve layer-specific effects. Recent 7T work by Horn et al. (2025) demonstrated layer-specific activity to noxious heat, highlighting the potential to distinguish fiber-specific projections within the GM with higher-resolution ultra-high field spinal imaging.

High lateralization to the right hemicord was found for Amp4 and for the linear contrast (**Figure 7**), consistent with stimulation of the right hand and with previous studies (Weber et al., 2016a, 2016b). We did not observe increasingly bilateral responses with higher intensity. This is in line with Brooks et al (Brooks et al., 2012) who reported ipsilateral responses for both thermal innocuous and noxious stimulation. However, this contrasts with studies reporting more bilateral spinal cord activity for noxious stimulation, such as Summers et al. (Summers et al., 2010) using noxious laser stimuli, and Weber et al. (Weber et al., 2016a) who found bilateral responses (along with ventral horn involvement and rostrocaudal spread from C4–C8) during both innocuous and painful heat stimulation of the C6 dermatome.

#### 4.2.2. Intensity encoding

When examining stimulation intensities separately, group-level analyses (randomise, Z 2.3, FWE p < 0.050) revealed no significant activity for Amp1, Amp2, or Amp3, (although Amp3 yielded significant clusters within run-1). In contrast, Amp4 produced significant activity localized to the right GM at the C7 spinal segment level

This pattern is consistent with the expectation that increasing stimulation amplitude increases effective afferent drive, thereby producing stronger BOLD responses. In our study, Amp4 was also frequently perceived as painful, suggesting that the Amp4 effect may reflect, at least in part, the transition from innocuous to noxious stimulation: 6/40 participants reported Amp1 as painful compared to 35/40 for Amp4. Consistent with this interpretation, Summers et al. (Summers et al., 2010) reported stronger spinal BOLD responses and more bilateral activity for noxious thermal stimulation compared with innocuous brushing.

### 4.3. Reliability across runs

We explored the reliability of the BOLD activity across runs and found that for Amp4, reliability at the C7 segment was moderate (ICC ≈ 0.5; **Figure 8**), noting that estimating ICC across three runs can yield lower values than pairwise comparisons. To understand this limited repeatability, we examined each run separately. At the participant level, the number of active voxels decreased significantly from run 1 to runs 2 and 3 across all segment levels within the right GM (**Figure 9**), whereas mean Z scores did not change. At the group level, run-1 produced significant clusters (Amp3, Amp4, and the linear contrast; **Figure S7**), but no FWE-corrected clusters were detected in runs 2 or 3 for any intensity. Importantly, we did not observe a comparable decline within run-1 across trials (**Figure S4**), suggesting that the effect emerged primarily between runs rather than within a run.

This decrease in signal across runs could reflect habituation or adaptation of the participant to the electrical stimulation. Although maximum pain ratings did not change significantly across runs, they trended downward (**Figure S6**); we did not investigate the perception of the stimulation intensity of the participant. Increased head/neck motion is an alternative explanation for reduced reliability across runs, but the number of motion outliers did not increase across runs (**Figure S5**).

Evidence for habituation to electrical stimulation in spinal cord fMRI is limited. Kündig et al. (2025) reported substantially reduced detected activity during a second session of lumbosacral spinal cord fMRI using non-noxious tibial nerve stimulation. Habituation to repeated electrical stimuli is also well documented in brain fMRI, particularly for noxious stimulation, where activity in pain-related regions decreases over repeated noxious electric stimuli (Nickel et al., 2014). Paul et al (2021) showed that habituation patterns differed for innocuous versus noxious stimulation of the hand, with more generalized somatosensory habituation compared to pain-specific regions.

Habituation likely contributed to the reduced run-to-run repeatability observed here, but its physiological origin remains uncertain. As electrocutaneous stimulation bypasses mechanoreceptors, peripheral receptor adaptation is unlikely. Adaptation of the peripheral nerve level is also improbable under the parameters used, as peripheral nerve fatigue typically requires higher-frequency stimulation (200 Hz) then was applied here (Graczyk et al., 2018). More plausible explanations include reduced excitability at the first synapse in the dorsal horn (e.g., presynaptic inhibition or other short-term plasticity mechanisms) and/or increased descending modulation from supraspinal regions, both of which have been implicated in sensory attenuation during repeated stimulation. Prior work also suggests that habituation can build over time, leading to progressively faster signal attenuation in subsequent runs, which is consistent with our observations (Higashiyama & Tashiro, 1989).

Several methodological factors may have accelerated habituation in this experiment, including testing four stimulation amplitudes within the same session and using a fixed inter-run rest interval. Higashiyama et al. (Higashiyama & Tashiro, 1989) investigated adaptation to electrocutaneous stimulation across levels spanning sensation to pain thresholds and reported greater adaptation at lower pulse amplitudes, with slower adaptation at higher amplitudes. In our study, because four amplitude levels were presented in randomized order, the lower amplitudes may have contributed to faster overall adaptation or habituation. Similar results were reported by Buma et al. (Buma et al., 2007) although adaptation there was quantified perceptually rather than using fMRI (Buma et al., 2007; Higashiyama & Tashiro, 1989).

Future studies may incorporate longer and/or randomized rest periods, consider limiting the number of amplitude conditions per session, and re-evaluate stimulation parameters such as pulse width, frequency, and waveform (monophasic versus biphasic; anodal versus cathodal) to reduce the effects of habituation (Herrero et al., 2000; A. Y. J. Szeto & Mao, 1982). For example, Szeto et al. (A. Y. Szeto & Saunders, 1982) reported greater adaptation at higher stimulation frequencies, and (Kaczmarek, 2000; A. Y. J. Szeto & Mao, 1982) found that 15–30 min rest periods allowed recovery from adaptation. In addition, while many neurophysiological protocols set intensity relative to the detection threshold, some paradigms increase intensity across repeated runs to counteract habituation, which may be worth testing in future spinal fMRI work (Nierula et al., 2024).

In contrast to the decrease in activity across runs observed here, Weber et al. (Weber et al., 2016a) reported evidence of sensitization in the spinal cord during painful thermal stimuli related by an increased spatial extent of the activity across runs, which was not the case with the non-painful heat stimulus.

### 4.4. Limitations and future work

This study has several limitations. First, we did not investigate sex and age effects. Although enrollment was balanced (20 females, 20 males), the study was not powered to detect sex differences. Our sample-size analysis indicated that n = 28 participants were required to reliably detect the C7 activity (**Figure S9**), leaving limited power for subgroup comparisons. Second, there is no standardized spinal fMRI preprocessing pipeline, and several methodological choices remain unsettled, including the use of spatial smoothing and whether to include white matter as a nuisance regressor (Grajauskas et al., 2019). Because the latter remains debated, its impact on group-level activity will be evaluated in future analyses. Third, the present study targeted a single dermatome. Mapping multiple dermatomes will be necessary to determine whether stimulation of adjacent sites produces cross-dermatomal effects (e.g., habituation) and to test the generalizability of our findings. Fourth, robust segment-level localization was most evident at higher stimulation intensities and in the initial run, with clear attenuation across repeated runs. This run-related reduction in detectability, consistent with habituation/adaptation and/or other session-dependent factors, could bias multi-run paradigms if not explicitly modeled or mitigated. Fifth, stimulation intensities were individualized and subjective ratings were limited, constraining inferences about dose–response relationships and precluding mechanistic conclusions (e.g., fiber-type recruitment or specific inhibitory/excitatory processes). Finally, electrocutaneous stimulation may not engage the spinal cord in the same way as touch or other sensory modalities, which also warrants further study.

In our next study, we will address these questions by examining sex effects, multiple dermatome sites, neural injury, and preprocessing steps in a larger cohort of healthy participants and in patients with cervical radiculopathy.

## 5. Conclusion

We characterized spinal cord sensory processing within the spinal cord with electrocutaneous stimulation during fMRI. Spinal activity increased with stimulation intensity and was primarily localized within the right dorsal horn at the expected C7 spinal segment level. Importantly, rootlet-based spatial normalization improved segmental alignment across participants, yielding greater spatial specificity and sensitivity than conventional disc-based normalization. Responses attenuated across repeated runs, consistent with habituation/adaptation that can reduce detectability in multi-run paradigms if not modeled. Together, these findings support the use of spinal fMRI to directly examine how sensory signals are organized and distributed within the human spinal cord, and provide a methodological foundation for more accurate localization of cervical sensory processing in basic research and translational research.

## Data Availability

The data will be made available on openneuro.org.

## 6. Acknowledgments and funding sources

Computing for this project was performed on the Sherlock high-performance computing cluster. We would like to thank Stanford University and the Stanford Research Computing Center for providing computational resources and support that contributed to these findings.

This study was supported by the National Institute of Neurological Disorders and Stroke (grant number R01NS133305, R61NS118651, and K24NS126781). The content is solely the responsibility of the authors and does not necessarily represent the official views of the National Institutes of Health. Funded by the Canada Research Chair in Quantitative Magnetic Resonance Imaging [CRC-2020-00179], the Canadian Institute of Health Research [PJT-190258], the Canada Foundation for Innovation [32454,34824], the Fonds de Recherche du Québec-Santé [322736,324636], the NSERC of Canada [RGPIN-2019-07244], the Canada First Research Excellence Fund (IVADO and TransMedTech), the Courtois NeuroMod project, the Quebec BioImaging Network [5886, 35450], INSPIRED, Mila-Tech Transfer Funding Program. SB is supported by the Canada Graduate Scholarships-Doctoral program from the NSERC of Canada. DP was supported by a Swiss National Science Foundation, Postdoc Mobility Fellowship grant (P500PM_214211) and the Redlich Pain Research Endowment.

## 7. Ethics

The study was approved by Stanford University’s Institutional Review Board.

## 8. Data and code availability

The code for preprocessing and data analysis is available on GitHub: https://github.com/kennethaweberii/Dermatomal_Mapping_R01/releases/tag/r20260213

The data will be made available on openneuro.org.

## 9. Author Contributions

Conceptualization: GG, KW; Data curation: SB, MK, TI, CL; Formal analysis: SB, MK, TI, CL, DP, LL, ZS; Funding acquisition: JR, SH, ST, SM, JCA, KW; Investigation: SB, MK, TI, CL, GHG, KW; Methodology: SB, MK, CL, KW; Project administration: KW Resources: GHG, JCA, KW; Software: SB, MK, JCA, KW; Supervision: MK, SM, JCA, KW; Validation: SB, MK; Visualization: SB, MK, LL; Writing – original draft: SB; Writing – review & editing: MK, TI, CL, DP, LL, JR, SH, ST, ZS, GG, SM, JCA, KW

## 10. Declaration of competing interests

The authors declared no potential conflicts of interest with respect to the research, authorship, and/or publication of this article.

During the preparation of this work the authors used ChatGPT-5.2 in order to assist with refining professional English. After using this tool, the authors reviewed and edited the content as needed and take full responsibility for the content of the published article.

## 12. Supplementary material

**Figure S1.**
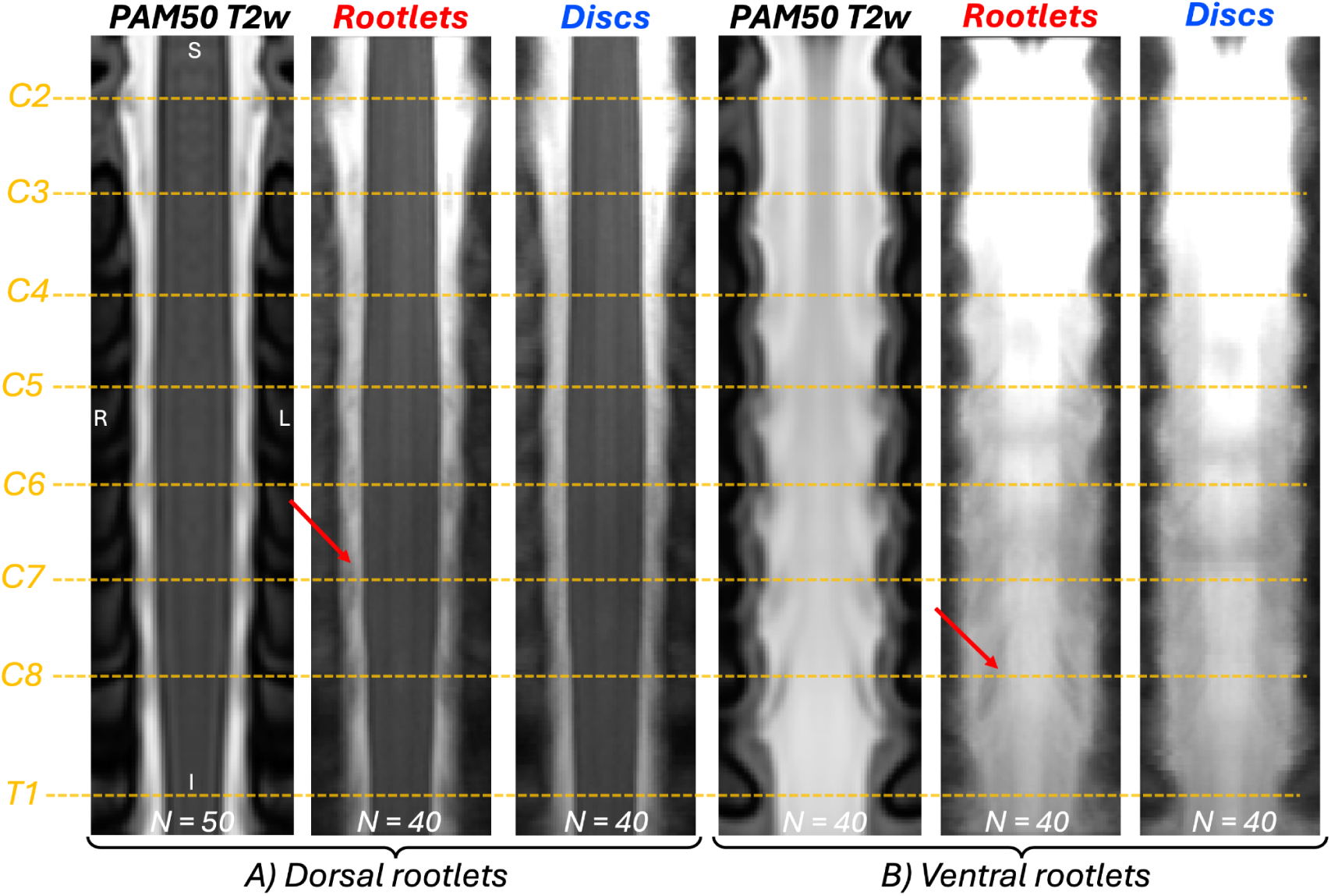
Group Mean T2w image (N = 40) registered to the PAM50 template using either rootlet-based (red) or disc-based (blue) spatial normalization. The PAM50 T2-weighted template image is shown for reference (black). Orange dashed lines mark the centers of spinal cord segments C2–T1 as defined in PAM50. **A)** Representative coronal slice (y = 70) illustrating dorsal rootlets. **B)** Representative coronal slice (y = 79) illustrating ventral rootlets. Red arrows indicate examples of well-delineated rootlets. Overall, rootlets are sharper and show better correspondence across participants with rootlet-based normalization than with disc-based normalization.

**Figure S2.**
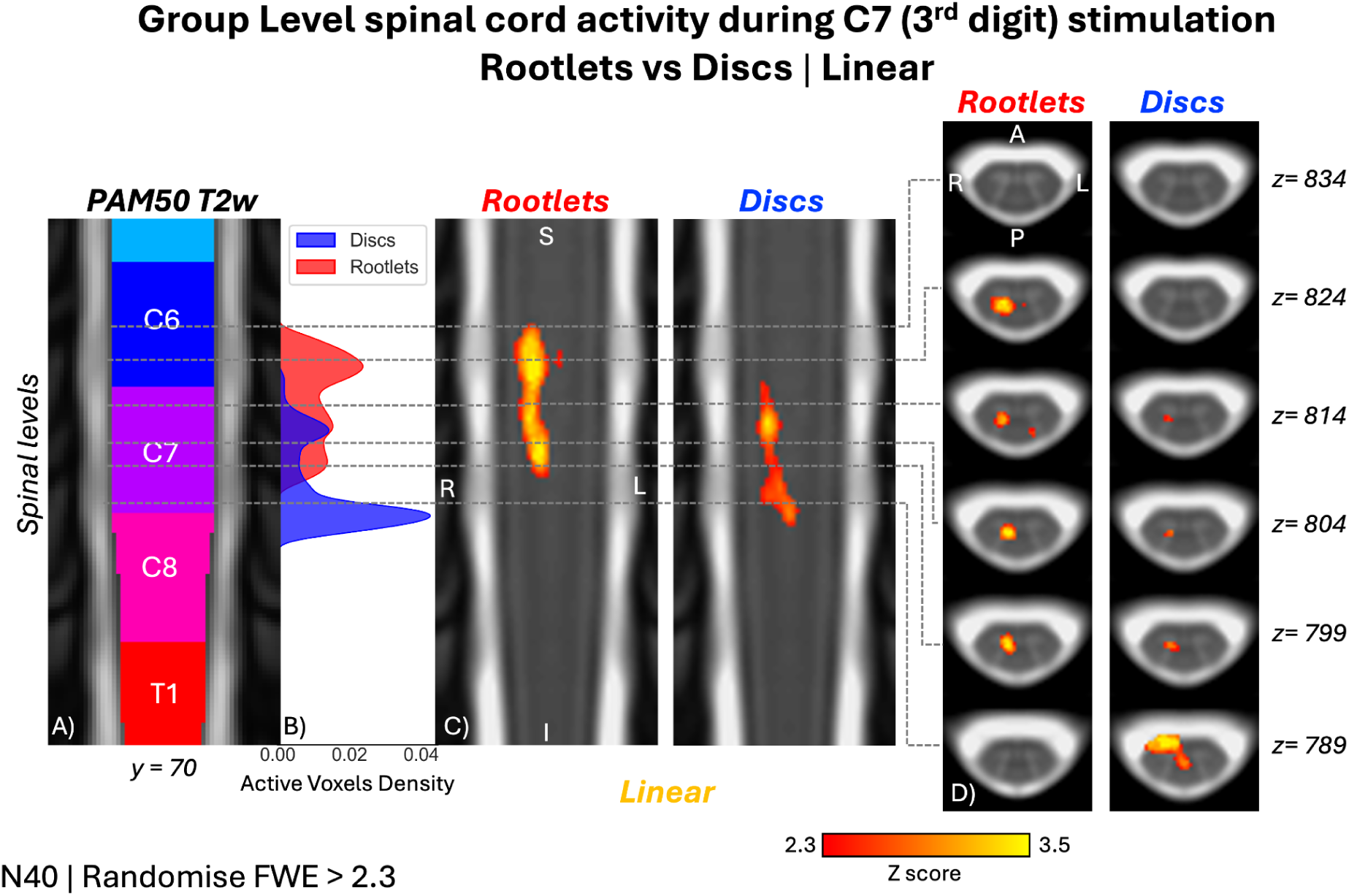
Group-level activity maps during electrocutaneous stimulation of the 3rd digit of the right hand (N = 40, Linear) comparing rootlets- (red) vs. discs-based (blue) normalization. **A)** Coronal view (y = 70, PAM50) with spinal segment levels. **B)** Active voxel density. **C)** Coronal slices with activity overlay. **D)** Axial views of representative clusters. Maps: Randomise, Z score > 2.3, and cluster corrected for family-wise error (FWE) at p < 0.050. In the coronal view, the cluster appears narrower and shows a higher Z score, whereas the disc shows less active voxels and lower Z score.

**Figure S3.**
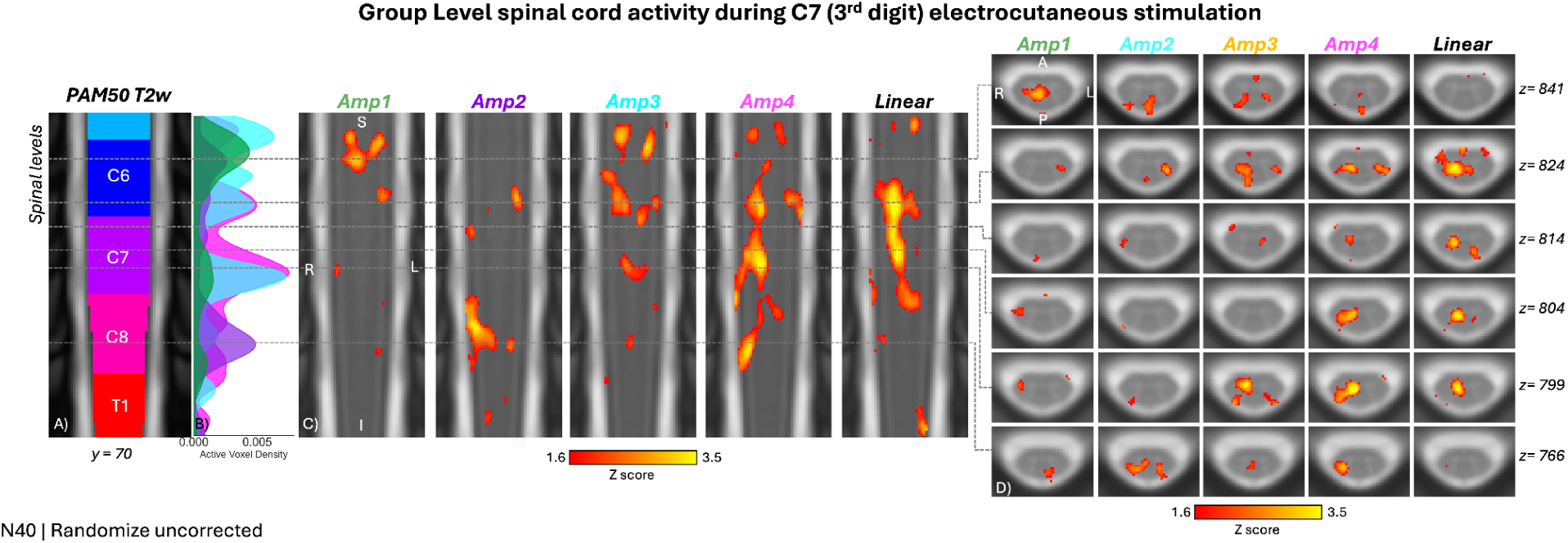
Group-level activity maps during electrocutaneous stimulation of the 3rd digit of the right hand (N = 40) with various stimulation amplitudes. **A)** Coronal view (y=70 on the PAM50 referential system) with an overlay of the spinal segment levels, matching the coverage of the functional EPI scans. **B)** Density plot of active voxels for stimulation amplitude Amp4 and linear fit between stimulation intensities. **C)** Coronal slices with an overlay of the activity maps. The location of the activity was assessed using the left-right (LR) ratio. **D)** Selected axial views centered on representative activity blobs. The activity maps were generated using a Randomise model, voxel-wise thresholded at Z score > 2.3, and uncorrected for family-wise-error.

**Figure S4.**
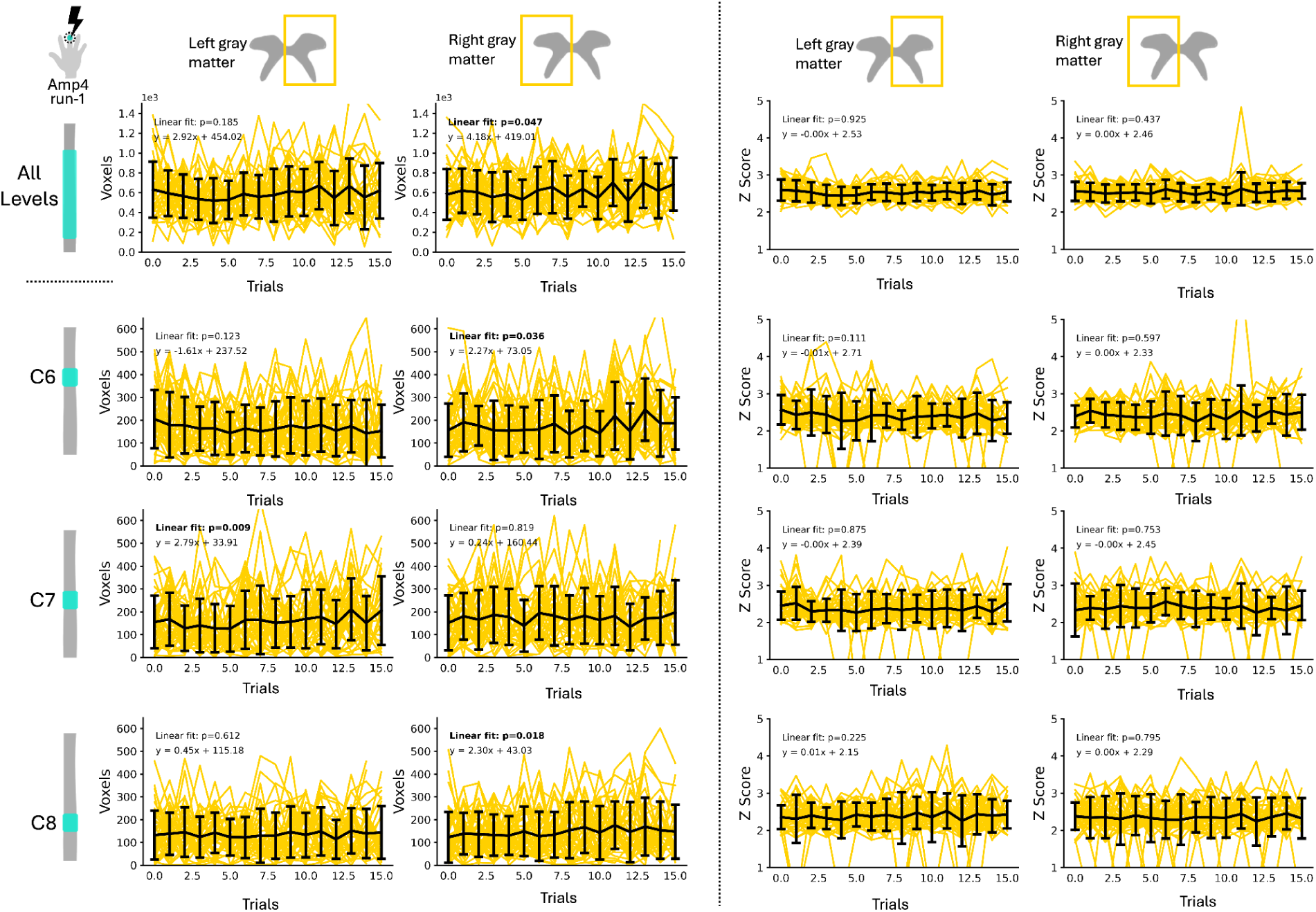
Average number of active voxels and Z score within left and right spinal cord gray matter within run 1 for intensity Amp4 for each stimulation trial (p < 0.050) for all spinal segment levels, C6, C7 and C8. Each yellow line represents one participant as the black line represents the average across all participants with the brackets showing the standard deviation. The number of active and average Z scores stay constant across trials. Linear fits are also shown.

**Figure S5.**
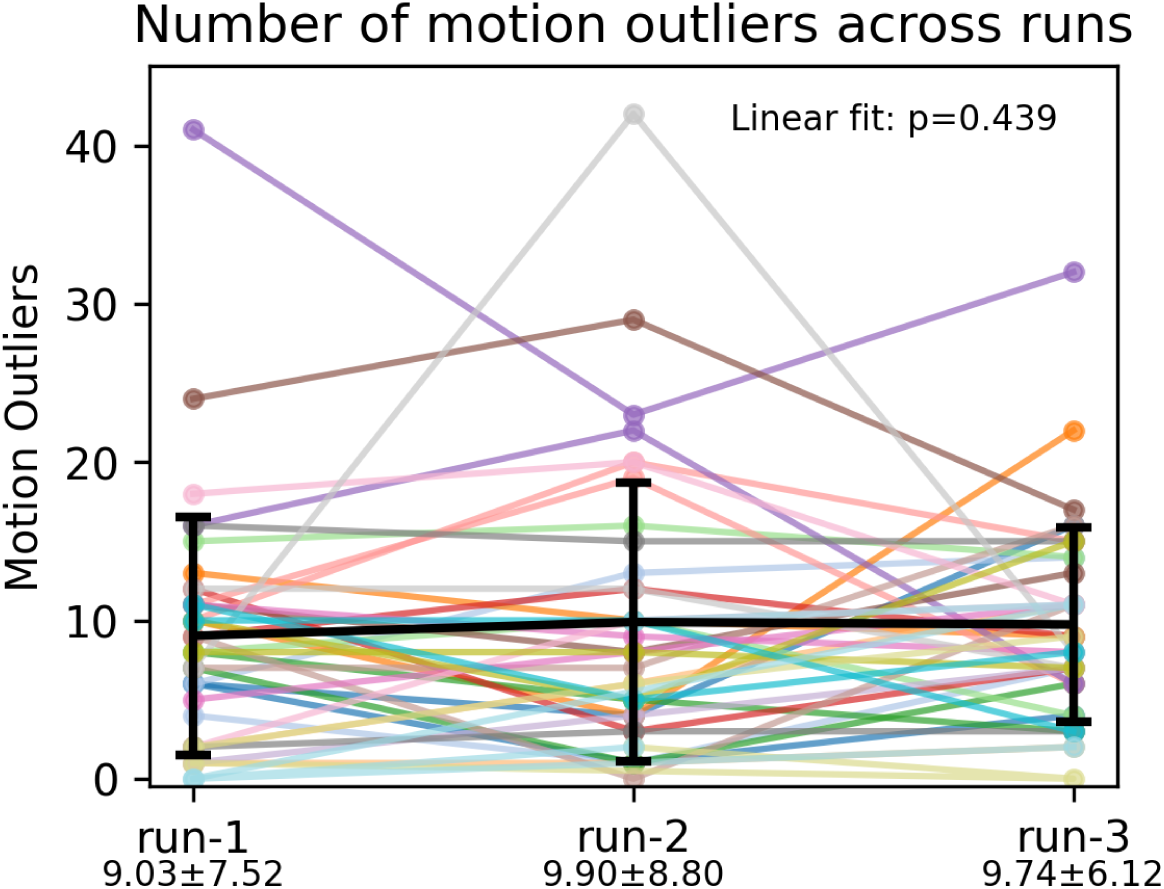
Number of motion outliers across participants and runs. Each line represents one participant as the black line represents the average across all participants with the brackets showing the standard deviation with values under each run (376 volumes per run). Motion outliers were measured based on DVARS which corresponds to the root mean square of the temporal change of the fMRI voxel-wise signal at each time point.

**Figure S6.**
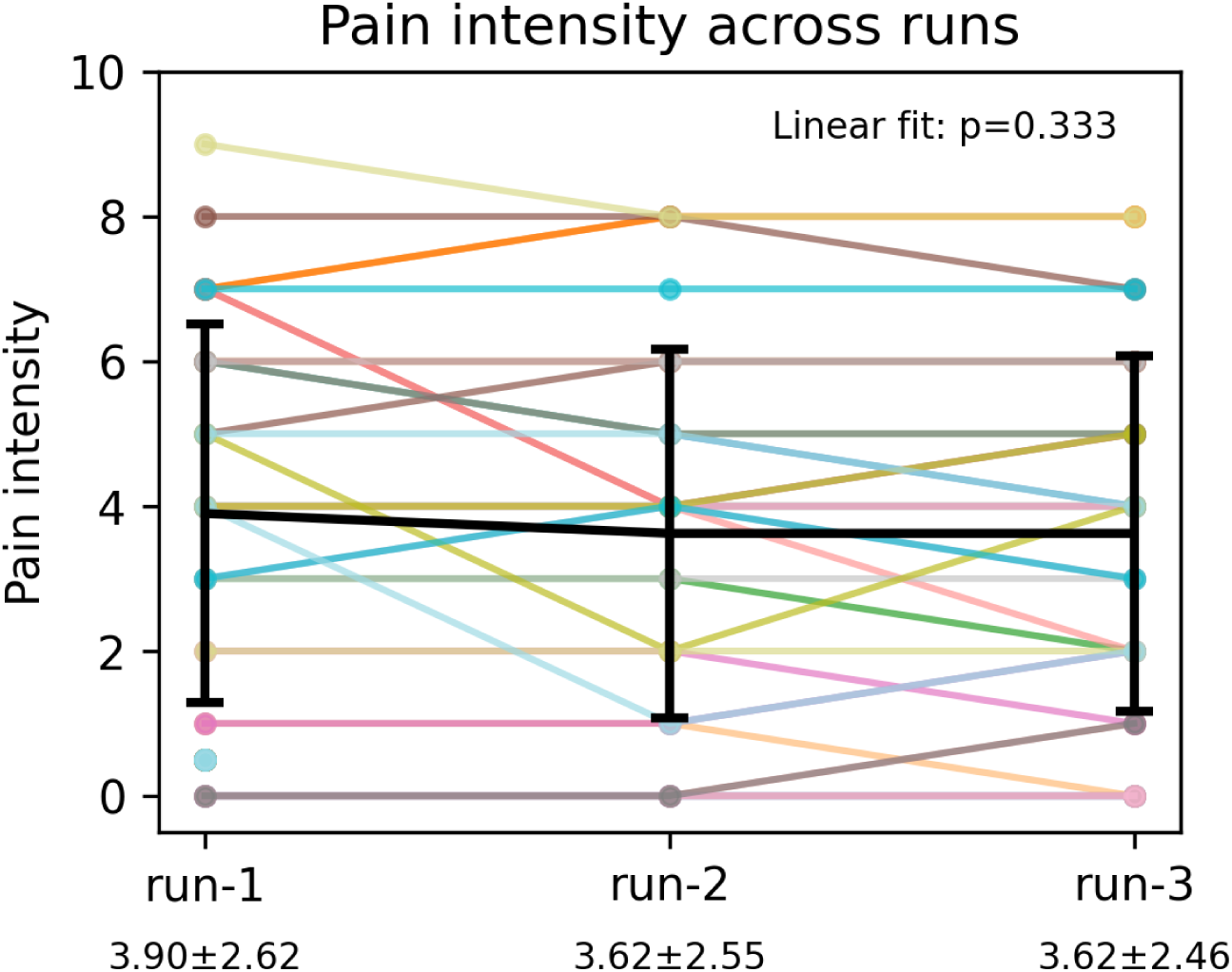
Maximum pain intensity across participants and runs. Each line represents one participant as the black line represents the average across all participants with the brackets showing the standard deviation with values under each run.

**Figure S7.**
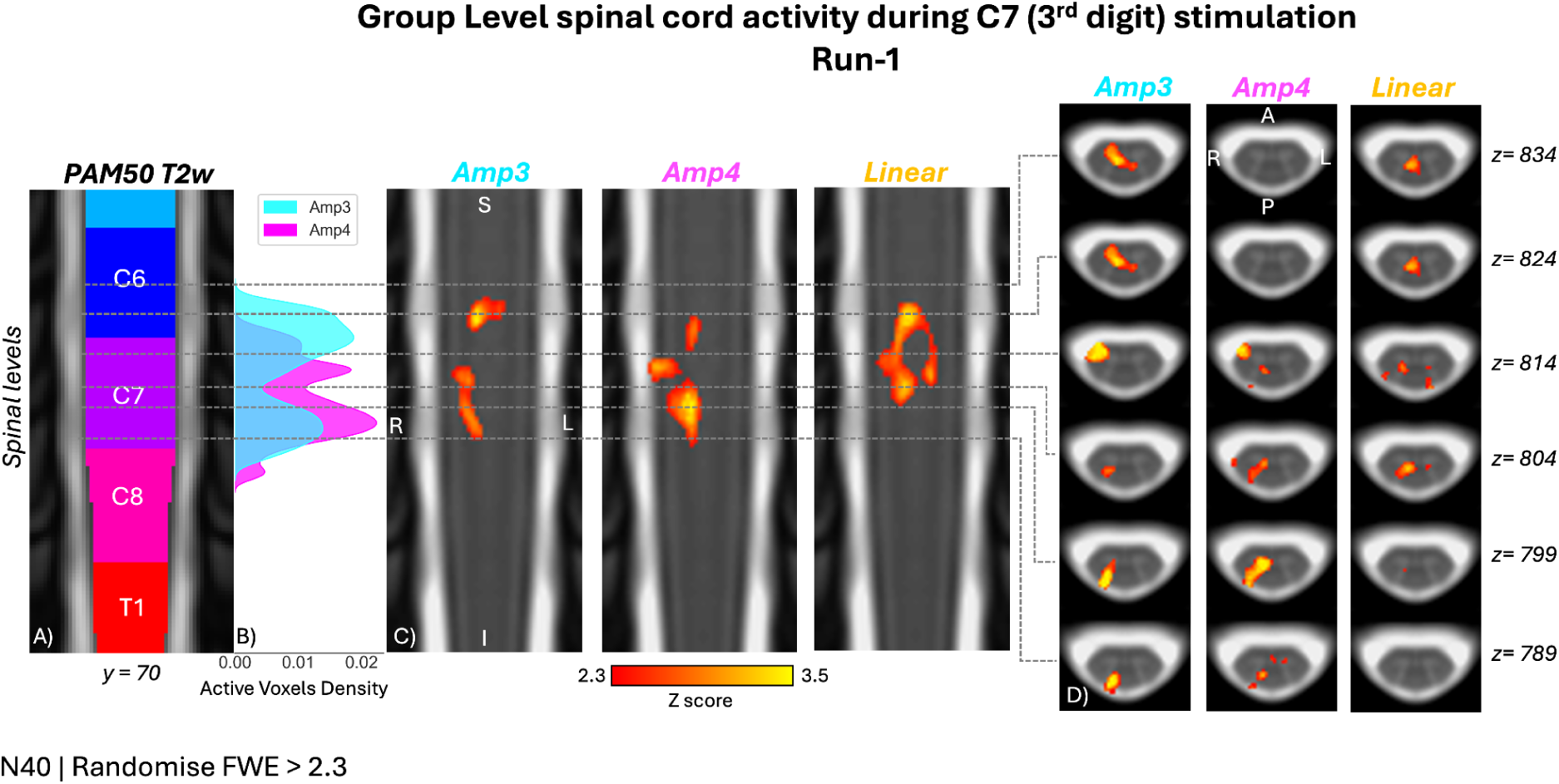
Run-1 group-level activity maps during electrocutaneous stimulation of the 3rd digit of the right hand (N = 40, Linear) for run-1. **A)** Coronal view (y=70, PAM50) with spinal segment levels. **B)** Active voxel density. **C)** Coronal slices with activity overlay. **D)** Axial views of representative clusters. Maps: Randomise, Z score > 2.3, and cluster corrected for family-wise error (FWE) at p < 0.050.

**Figure S8.**
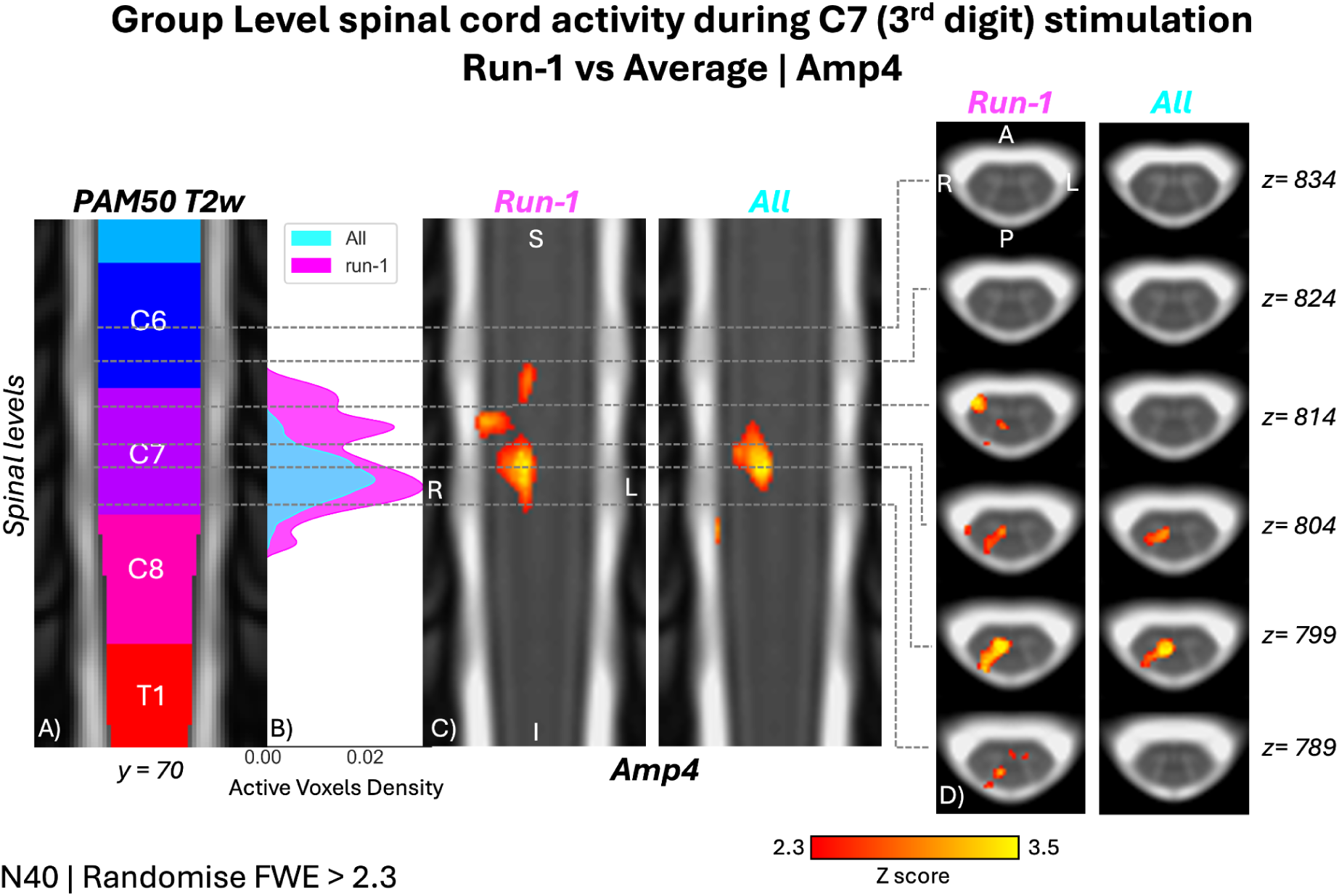
Run-1 and average of runs group-level activity maps during electrocutaneous stimulation of the 3rd digit of the right hand (N = 40, Linear) comparing run-1 only vs. average of 3 runs. **A)** Coronal view (y=70, PAM50) with spinal segment levels. **B)** Active voxel density. **C)** Coronal slices with activity overlay. **D)** Axial views of representative clusters. Maps: Randomise, Z score > 2.3, and cluster corrected for family-wise error (FWE) at p < 0.050.

### 12.1. How many participants are needed for a spinal cord fMRI study?

Figure S9 illustrates the voxelwise detection frequency across 1,000 Monte Carlo iterations for Amp4 (averaged across three runs) as a function of sample size (Randomise with Z > 2.3 and FWE cluster correction p < 0.050). As the sample size increases, the cluster emerging at the C7 spinal segment level becomes more stable. Detection frequency surpasses 50% at approximately N ≈ 28, and larger cohorts yield more focal and reliable activity patterns that converge toward the full-cohort (N = 40) group level result.

**Figure S9.**
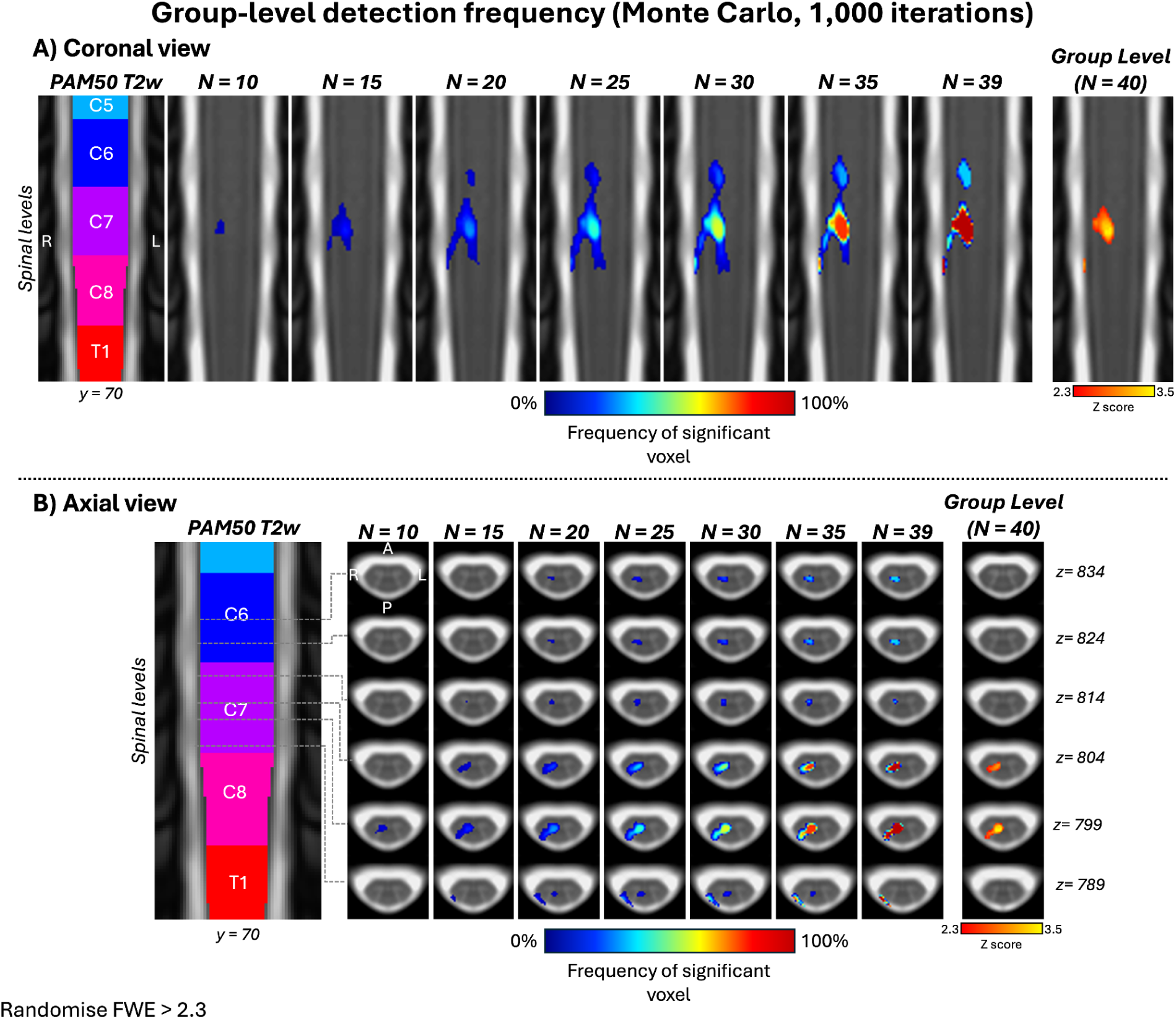
Group level detection frequency map across sample sizes. **A)** Coronal and **B)** axial views showing voxelwise detection frequency maps derived from group-level analyses using Randomise with Z > 2.3 and family-wise error (FWE) cluster correction. activity corresponds to the Amp4 stimulation, averaged across three runs. For each sample size (N = 10, 15, 20, 25, 30, 35, 39), subsamples were drawn without replacement, and 1,000 Monte-Carlo iterations were performed to compute the percentage of iterations in which each voxel was significant. The rightmost column shows the corresponding full group-level map (N = 40) for comparison.

## References

Banerjee, R., Kaptan, M., Tinnermann, A., Khatibi, A., Dabbagh, A., Büchel, C., Kündig, C. W., Law, C. S. W., Pfyffer, D., Lythgoe, D. J., Tsivaka, D., Van De Ville, D., Eippert, F., Muhammad, F., Glover, G. H., David, G., Haynes, G., Haaker, J., Brooks, J. C. W., … Cohen-Adad, J. (2025). EPISeg: Automated segmentation of the spinal cord on echo planar images using open-access multi-center data. In bioRxivorg. 10.1101/2025.01.07.631402

Bédard, S., Karthik, E. N., Tsagkas, C., Pravatà, E., Granziera, C., Smith, A., Weber, K. A., II, & Cohen-Adad, J. (2025). Towards contrast-agnostic soft segmentation of the spinal cord. Medical Image Analysis, 103473, 103473. 10.1016/j.media.2025.103473

Bédard, S., Valošek, J., Oliva, V., Weber, K. A., Ii, & Cohen-Adad, J. (2025). Rootlets-based registration to the PAM50 spinal cord template. Imaging Neuroscience (Cambridge, Mass.), 3(IMAG.a.123). 10.1162/IMAG.a.123

Brooks, J. C. W., Beckmann, C. F., Miller, K. L., Wise, R. G., Porro, C. A., Tracey, I., & Jenkinson, M. (2008). Physiological noise modelling for spinal functional magnetic resonance imaging studies. NeuroImage, 39(2), 680–692. 10.1016/j.neuroimage.2007.09.018

Brooks, J. C. W., Kong, Y., Lee, M. C., Warnaby, C. E., Wanigasekera, V., Jenkinson, M., & Tracey, I. (2012). Stimulus site and modality dependence of functional activity within the human spinal cord. The Journal of Neuroscience: The Official Journal of the Society for Neuroscience, 32(18), 6231–6239. 10.1523/JNEUROSCI.2543-11.2012

Buma, D. G., Buitenweg, J. R., & Veltink, P. H. (2007). Intermittent stimulation delays adaptation to electrocutaneous sensory feedback. IEEE Transactions on Neural Systems and Rehabilitation Engineering, 15(3), 435–441. 10.1109/TNSRE.2007.903942

Caceres, A., Hall, D. L., Zelaya, F. O., Williams, S. C. R., & Mehta, M. A. (2009). Measuring fMRI reliability with the intra-class correlation coefficient. NeuroImage, 45(3), 758–768. 10.1016/j.neuroimage.2008.12.035

Cadotte, D. W., Cadotte, A., Cohen-Adad, J., Fleet, D., Livne, M., Wilson, J. R., Mikulis, D., Nugaeva, N., & Fehlings, M. G. (2015). Characterizing the Location of Spinal and Vertebral Levels in the Human Cervical Spinal Cord. AJNR. American Journal of Neuroradiology, 36(4), 803–810. 10.3174/ajnr.A4192

Chung, K., & Coggeshall, R. E. (1983). Propriospinal fibers in the rat. The Journal of Comparative Neurology, 217(1), 47–53. 10.1002/cne.902170105

Cicchetti, D. V. (1994). Guidelines, criteria, and rules of thumb for evaluating normed and standardized assessment instruments in psychology. Psychological Assessment, 6(4), 284–290. 10.1037/1040-3590.6.4.284

Coggeshall, R. E., Chung, K., Chung, J. M., & Langford, L. A. (1981). Primary afferent axons in the tract of Lissauer in the monkey. The Journal of Comparative Neurology, 196(3), 431–442. 10.1002/cne.901960307

Cohen-Adad, J., Alonso-Ortiz, E., Abramovic, M., Arneitz, C., Atcheson, N., Barlow, L., Barry, R. L., Barth, M., Battiston, M., Büchel, C., Budde, M., Callot, V., Combes, A. J. E., De Leener, B., Descoteaux, M., de Sousa, P. L., Dostál, M., Doyon, J., Dvorak, A., … Xu, J. (2021). Generic acquisition protocol for quantitative MRI of the spinal cord. Nature Protocols, 16(10), 4611–4632. 10.1038/s41596-021-00588-0

De Leener, B., Dahlberg, S., Khatibi, L., Cohen-Adad, J., & Doyon, J. (2020). Effect non-protonated perfluorocarbon liquid-filled SatPads spinal cord MR imaging. ISMRM Annual Meeting Proceedings.

De Leener, B., Fonov, V. S., Collins, D. L., Callot, V., Stikov, N., & Cohen-Adad, J. (2018). PAM50: Unbiased multimodal template of the brainstem and spinal cord aligned with the ICBM152 space. NeuroImage, 165, 170–179. 10.1016/j.neuroimage.2017.10.041

De Leener, B., Kadoury, S., & Cohen-Adad, J. (2014). Robust, accurate and fast automatic segmentation of the spinal cord. NeuroImage, 98, 528–536. 10.1016/j.neuroimage.2014.04.051

De Leener, B., Lévy, S., Dupont, S. M., Fonov, V. S., Stikov, N., Louis Collins, D., Callot, V., & Cohen-Adad, J. (2017). SCT: Spinal Cord Toolbox, an open-source software for processing spinal cord MRI data. NeuroImage, 145(Pt A), 24–43. 10.1016/j.neuroimage.2016.10.009

Finsterbusch, J. (2014). B0 Inhomogeneity and Shimming. In Quantitative MRI of the Spinal Cord (pp. 68–90). Elsevier. 10.1016/b978-0-12-396973-6.00006-x

Graczyk, E. L., Delhaye, B. P., Schiefer, M. A., Bensmaia, S. J., & Tyler, D. J. (2018). Sensory adaptation to electrical stimulation of the somatosensory nerves. Journal of Neural Engineering, 15(4), 046002. 10.1088/1741-2552/aab790

Grajauskas, L. A., Frizzell, T., Song, X., & D’Arcy, R. C. N. (2019). White matter fMRI activation cannot be treated as a nuisance regressor: Overcoming a historical blind spot. Frontiers in Neuroscience, 13, 1024. 10.3389/fnins.2019.01024

Herrero, J. F., Laird, J. M., & López-García, J. A. (2000). Wind-up of spinal cord neurones and pain sensation: much ado about something? Progress in Neurobiology, 61(2), 169–203. 10.1016/s0301-0082(99)00051-9

Higashiyama, A., & Tashiro, T. (1989). Magnitude estimates for electrical pulses: evidence for two neural mechanisms. Perception & Psychophysics, 45(6), 537–549. 10.3758/bf03208061

Horn, U., Vannesjo, S. J., Gross-Weege, N., Trampel, R., Revina, Y., Kaptan, M., Dabbagh, A., Beghini, L., Callot, V., Todd, A., Pine, K. J., Moeller, H. E., Finsterbusch, J., Weiskopf, N., & Eippert, F. (2025). Ultra-high-field fMRI reveals layer-specific responses in the human spinal cord. In bioRxiv. 10.1101/2025.07.17.665316

Islam, H., Law, C. S. W., Weber, K. A., Mackey, S. C., & Glover, G. H. (2019). Dynamic per slice shimming for simultaneous brain and spinal cord fMRI. Magnetic Resonance in Medicine: Official Journal of the Society of Magnetic Resonance in Medicine / Society of Magnetic Resonance in Medicine, 81(2), 825–838. 10.1002/mrm.27388

Jenkinson, M., Beckmann, C. F., Behrens, T. E. J., Woolrich, M. W., & Smith, S. M. (2012). FSL. NeuroImage, 62(2), 782–790. 10.1016/j.neuroimage.2011.09.015

Kaczmarek, K. A. (2000). Electrotactile adaptation on the abdomen: preliminary results. IEEE Transactions on Rehabilitation Engineering, 8(4), 499–505. 10.1109/86.895953

Karthik, E. N., Bédard, S., Valošek, J., Aigner, C. S., Bannier, E., Bednařík, J., Callot, V., Combes, A., Curt, A., David, G., Eippert, F., Farner, L., Fehlings, M. G., Freund, P., Granberg, T., Granziera, C., RHSCIR Network Imaging Group, Horn, U., Horák, T., … Cohen-Adad, J. (2025). Monitoring morphometric drift in lifelong learning segmentation of the spinal cord. In arXiv [cs.CV]. arXiv. http://arxiv.org/abs/2505.01364

Kinany, N., Pirondini, E., Micera, S., & Van De Ville, D. (2022). Spinal Cord fMRI: A New Window into the Central Nervous System. The Neuroscientist: A Review Journal Bringing Neurobiology, Neurology and Psychiatry, 10738584221101827. 10.1177/10738584221101827

Krejci, K., Chmelik, J., Bédard, S., Eippert, F., Horn, U., Callot, V., Cohen-Adad, J., & Valosek, J. (2025). RootletSeg: Deep learning method for spinal rootlets segmentation across MRI contrasts. In arXiv [q-bio.TO]. arXiv. http://arxiv.org/abs/2509.16255

Kündig, C., Büeler, S., Liechti, M., Kessler, T., Freund, P., & David, G. (2025). Repeated functional magnetic resonance imaging of the lumbosacral cord during electrical stimulation of the tibial nerve. ISMRM Annual Meeting, Article 0200. 2025 ISMRM & ISMRT Annual Meeting, Honolulu, Hawaii, USA. 10.58530/2025/0200

Landelle, C., Lungu, O., Vahdat, S., Kavounoudias, A., Marchand-Pauvert, V., De Leener, B., & Doyon, J. (2021). Investigating the human spinal sensorimotor pathways through functional magnetic resonance imaging. NeuroImage, 245(118684), 118684. 10.1016/j.neuroimage.2021.118684

Lawrence, J. M., Stroman, P. W., & Kollias, S. S. (2008). Functional magnetic resonance imaging of the human spinal cord during vibration stimulation of different dermatomes. Neuroradiology, 50(3), 273–280. 10.1007/s00234-007-0338-6

Lee, M. W. L., McPhee, R. W., & Stringer, M. D. (2008). An evidence-based approach to human dermatomes. *Clinical Anatomy (New York*, N.Y.), 21(5), 363–373. 10.1002/ca.20636

Light, A. R., & Perl, E. R. (1979). Spinal termination of functionally identified primary afferent neurons with slowly conducting myelinated fibers. The Journal of Comparative Neurology, 186(2), 133–150. 10.1002/cne.901860203

Lissarrague, M., Miranda Solís, F., & Martínez Benia, F. (2025). Intradural anastomoses between the cervical spinal nerves: Anatomical study. Neurocirugía (English Edition), 36(5), 500666. 10.1016/j.neucie.2025.500666

Moriishi, J., Otani, K., Tanaka, K., & Inoue, S. (1989). The intersegmental anastomoses between spinal nerve roots. The Anatomical Record, 224(1), 110–116. 10.1002/ar.1092240114

Nathan, P. W., & Smith, M. C. (1959). Fasciculi proprii of the spinal cord in man: Review of present knowledge. Brain: A Journal of Neurology, 82(4), 610–668. 10.1093/brain/82.4.610

Nickel, F. T., Ott, S., Möhringer, S., Saake, M., Dörfler, A., Seifert, F., & Maihöfner, C. (2014). Brain correlates of short-term habituation to repetitive electrical noxious stimulation. *European Journal of Pain (London*, England), 18(1), 56–66. 10.1002/j.1532-2149.2013.00339.x

Nierula, B., Stephani, T., Bailey, E., Kaptan, M., Pohle, L.-M. G., Horn, U., Mouraux, A., Maess, B., Villringer, A., Curio, G., Nikulin, V. V., & Eippert, F. (2024). A multichannel electrophysiological approach to noninvasively and precisely record human spinal cord activity. PLoS Biology, 22(10), e3002828. 10.1371/journal.pbio.3002828

Paul, K., Tik, M., Hahn, A., Sladky, R., Geissberger, N., Wirth, E.-M., Kranz, G. S., Pfabigan, D. M., Kraus, C., Lanzenberger, R., Lamm, C., & Windischberger, C. (2021). Give me a pain that I am used to: distinct habituation patterns to painful and non-painful stimulation. Scientific Reports, 11(1), 22929. 10.1038/s41598-021-01881-4

Pierrot-Deseilligny, E., & Marchand-Pauvert, V. (2002). A cervical propriospinal system in man. Advances in Experimental Medicine and Biology, 508, 273–279. 10.1007/978-1-4615-0713-0_33

Soteropoulos, D. S., Edgley, S. A., & Baker, S. N. (2013). Spinal commissural connections to motoneurons controlling the primate hand and wrist. The Journal of Neuroscience: The Official Journal of the Society for Neuroscience, 33(23), 9614–9625. 10.1523/JNEUROSCI.0269-13.2013

Stenner, M.-P., Nossa, C. M., Zaehle, T., Azañón, E., Heinze, H.-J., Deliano, M., & Büntjen, L. (2025). Prior knowledge changes initial sensory processing in the human spinal cord. Science Advances, 11(3), eadl5602. 10.1126/sciadv.adl5602

Stracke, C. P., Pettersson, L. G., Schoth, F., Möller-Hartmann, W., & Krings, T. (2005). Interneuronal systems of the cervical spinal cord assessed with BOLD imaging at 1.5 T. Neuroradiology, 47(2), 127–133. 10.1007/s00234-004-1318-8

Sugiura, Y., Lee, C. L., & Perl, E. R. (1986). Central projections of identified, unmyelinated (C) afferent fibers innervating mammalian skin. Science (New York, N.Y.), 234(4774), 358–361. 10.1126/science.3764416

Summers, P. E., Ferraro, D., Duzzi, D., Lui, F., Iannetti, G. D., & Porro, C. A. (2010). A quantitative comparison of BOLD fMRI responses to noxious and innocuous stimuli in the human spinal cord. NeuroImage, 50(4), 1408–1415. 10.1016/j.neuroimage.2010.01.043

Szeto, A. Y. J., & Mao, L. (1982). Dermal effects of electrocutaneous stimulation. In S. Saha (Ed.), Biomedical Engineering I (pp. 121–124). Elsevier. 10.1016/b978-0-08-028826-0.50030-5

Szeto, A. Y., & Saunders, F. A. (1982). Electrocutaneous stimulation for sensory communication in rehabilitation engineering. IEEE Transactions on Bio-Medical Engineering, 29(4), 300–308. https://pubmed.ncbi.nlm.nih.gov/7068167/

Valošek, J., & Cohen-Adad, J. (2024). Reproducible Spinal Cord Quantitative MRI Analysis with the Spinal Cord Toolbox. rev. 2023–0159. https://scholar.google.com/citations?view_op=view_citation&hl=en&citation_for_view=ZRId5X8AAAAJ:KlAtU1dfN6UC

Valošek, J., Mathieu, T., Schlienger, R., Kowalczyk, O. S., & Cohen-Adad, J. (2024). Automatic segmentation of the spinal cord nerve rootlets. Imaging Neuroscience, 2, 1–14. 10.1162/imag_a_00218

Weber, K. A., 2nd, Chen, Y., Paliwal, M., Law, C. S., Hopkins, B. S., Mackey, S., Dhaher, Y., Parrish, T. B., & Smith, Z. A. (2020). Assessing the spatial distribution of cervical spinal cord activity during tactile stimulation of the upper extremity in humans with functional magnetic resonance imaging. NeuroImage, 217(116905), 116905. 10.1016/j.neuroimage.2020.116905

Weber, K. A., 2nd, Chen, Y., Wang, X., Kahnt, T., & Parrish, T. B. (2016a). Functional magnetic resonance imaging of the cervical spinal cord during thermal stimulation across consecutive runs. NeuroImage, 143, 267–279. 10.1016/j.neuroimage.2016.09.015

Weber, K. A., 2nd, Chen, Y., Wang, X., Kahnt, T., & Parrish, T. B. (2016b). Lateralization of cervical spinal cord activity during an isometric upper extremity motor task with functional magnetic resonance imaging. NeuroImage, 125, 233–243. 10.1016/j.neuroimage.2015.10.014

Weber, K. A., Chen, Y., Wang, X., & Parrish, T. B. (2014). Choice of motion correction method affects spinal cord fMRI results. 20th Annual Meeting of the Organization for Human Brain Mapping.

Willis, W. D., & Westlund, K. N. (1997). Neuroanatomy of the pain system and of the pathways that modulate pain. Journal of Clinical Neurophysiology: Official Publication of the American Electroencephalographic Society, 14(1), 2–31. 10.1097/00004691-199701000-00002

Winkler, A. M., Ridgway, G. R., Webster, M. A., Smith, S. M., & Nichols, T. E. (2014). Permutation inference for the general linear model. NeuroImage, 92, 381–397. 10.1016/j.neuroimage.2014.01.060

